# A Systematic Review of Mathematical Models for Soil-Transmitted Helminthiasis Control Strategies

**DOI:** 10.1101/2025.08.05.25333090

**Authors:** Lemjini Masandawa, Miracle Amadi, Isambi S. Mbalawata, Safari Kinung’hi, Silas S. Mirau

## Abstract

**Background:** Soil-transmitted helminthiasis (STH) is a parasitic disease that affects over 1.5 billion people worldwide. The use of mathematical models to inform time-bound projections and support WHO targets is growing. Nevertheless, there is a lack of comprehensive synthesis regarding the extent to which mathematical frameworks for STH control account for the timeframe and effectiveness of interventions required to meet WHO programmatic goals.

**Objective:** The present study aims to bridge this gap by classifying existing mathematical models based on their structural design and population features, assessing the effectiveness and impact of interventions in achieving WHO targets, and examining the timelines specified in these models for attaining WHO morbidity control and elimination goals.

**Methods:** An extensive search was conducted in the Web of Science, Scopus, and Embase databases to retrieve articles published in English between January 2015 and December 2024. The search query captured terms related to mathematical models, disease, and interventions. Articles on mathematical models of STH transmission dynamics that evaluated at least one control strategy were included. Extracted data included the type of model, intervention, study characteristics, population structure model, WHO policy alignment, and predictive outcomes. A narrative synthesis was conducted, and results were summarized thematically. The quality of the published articles was assessed using the Assessment Modeling Studies Tool. The protocol for this review is registered at PROSPERO (CRD420251090375).

**Principal findings:** A total of 43 studies met the inclusion criteria. Stochastic models (47%) slightly outnumbered deterministic models (44%) and hybrid models (9%). Regarding model classification by population stratification, age-structured models (56%) were more prevalent compared to flat (28%) and spatial models (16%). Community- and school-based deworming were the most frequently modeled and compared interventions (42%), with community-based deworming (CBD) shown to interrupt STH transmission even in high-prevalence settings, outperforming child-targeted treatment strategies. Combination therapy was underexplored, representing only 7% of the studies. Sixty percent of the modeling studies aligned with the 2011–2020 morbidity control goals, while 40% aligned with the 2021–2030 transmission interruption objectives. Only 35% of the studies reported time-bound predictions, with significant variation in the duration required for a modeled intervention to reach WHO goals. The projected duration for attaining WHO targets ranged from 2 to 35 years under different intervention scenarios.

**Conclusions/significance:** The findings highlight the dominance of age-structured and stochastic models. Additionally, models demonstrate that community-wide treatment consistently outperforms child-targeted treatment strategies across most settings. However, there is insufficient time-bound modeling, underscoring the need for future research to explicitly include timelines for reaching WHO targets.

**Author summary:** Soil-transmitted helminthiases are parasitic worm infections acquired through contact with contaminated soil, primarily in low-resource settings. The World Health Organization (WHO) has established guidelines aimed at reducing the disease burden through either morbidity control or interruption of transmission. Mathematical models are essential tools for designing and evaluating STH control strategies in relation to meeting WHO-set targets. Many existing models report on reductions in prevalence or infection intensity. However, despite alignment with WHO goals, the time horizon required for modeled interventions to achieve the recommended targets has not been systematically synthesized. Therefore, this review synthesizes existing models by examining their structural design and population representation, how they predict the outcomes of interventions, and the timelines projected to achieve WHO goals.

In this study, we synthesized 43 modeling studies and found that stochastic models (47%) were slightly more common than deterministic (44%) and hybrid (9%) approaches. Age-structured models (56%) were more widely used than spatial (16%) and non-structured (28%) frameworks. Comparisons between school-based deworming (SBD) and community-based deworming (CBD) (42%) were among the most frequently modeled interventions. Although all 43 studies aligned with WHO targets, only a minority (35%) explicitly reported the time required for modeled interventions to achieve morbidity control or elimination goals.

Our findings highlight a critical gap: the lack of time-bound predictions in STH modeling. Addressing this gap in future research could improve strategic planning and policy decisions aimed at eliminating these infections.

## 1 Introduction

Soil-transmitted helminthiasis (STH) is among of the disease infections categorized by the World Health Organization (WHO) as neglected tropical diseases (NTDs), due to the fact that they receive little attention in research investments [42]. It is an intestinal infection primarily caused by three species: *Ascaris lumbricoides*, *Trichuris trichiura*, and hookworms (*Necator americanus* and *Ancylostoma duodenale*) [42, 43, 38]. STH affects more than 1.5 billion people in around 166 endemic countries globally [47, 53, 54]. Furthermore, the literature reveals that 4.5 billion people worldwide are estimated to be at risk, with associated disease burden ranging from 5 to 39 million disability-adjusted life years (DALYs) [30]. STH predominantly affects impoverished and marginalized communities who have difficulty in accessing safe drinking water, sufficient sanitation infrastructure, and proper hand-washing habits particularly Sub-Saharan Africa, China, Latin America, and South-East Asia [40].

Heavy infections with STH reveal clinical manifestations such as fatigue, tiredness, abdominal pain, diarrhea, chronic intestinal blood loss, skin rashes, dry hair, and loss of appetite. The said health consequences may exacerbates malnutrition and increase the rate of iron deficiency anemia [46]. Moreover, a high prevalence of STH is associated with disease morbidity such as impaired physical development (loss in weight and height), educational deficits (poor performance in school, absenteeism, reduced IQ), and reduced work productivity in adults (due to illness or absenteeism). STH morbidity perpetuates the cycle of poverty due to healthcare costs and social stigma [33]. The extent and severity of the burden imposed by STH in the human population is measured in terms of years of life lost due to premature death and years lived with disabilities, leading to disability-adjusted life years (DALYs) [54].

In response to the burden of STH, the World Health Organization (WHO) created two roadmaps in quick succession that outlined the global goals and tactics required to achieve them [28]. The first WHO roadmap (2011–2020) aimed at morbidity control in school-aged children (SAC) by regularly administering preventive chemotherapy in endemic settings. Later, the strategy was expanded to include pre-school-aged children (PSAC), pregnant women, and other at-risk groups. Despite these efforts, STH continued to persist due to high reinfection risks in some settings, necessitating further modification of strategies in terms of coverage regimens.

The current WHO’s roadmap (2021–2030) focuses on interrupting STH transmission or eliminating the disease as a public health problem in some settings [49]. The transition from the first to the second roadmap emphasizes more aggressive and targeted control measures for at-risk groups or communities, depending on resource availability. To meet the goal, the roadmap calls for supplementary interventions to preventive chemotherapy (PC), such as water, sanitation, and hygiene (WASH), behavioral interventions, improved detection tools such as qPCR, and strengthened monitoring systems.

Mathematical models are increasingly employed to design, refine, and evaluate control strategies [36, 37]. These models are essential for understanding the dynamics of disease transmission [59, 58], assessing the impact and efficacy of interventions, allocating resources as efficiently as possible, and evaluating both short- and long-term results based on different hypotheses. The timeframes and coverage level needed to reach global health goals can also be estimated using models. Deterministic, stochastic, and hybrid models are among the modeling techniques used to capture the homogeneity or heterogeneity of populations in terms of age and location. These models vary depending on structural assumptions, population stratification, and data requirements. Importantly, mathematical models guide policymakers in making decisions on specific control strategies and evaluating timelines for morbidity control, disease interruption, or elimination. It is crucial to consolidate these dispersed studies from the literature, even with the rise in distinct mathematical methodologies. Thus, the current study provides a summary of how mathematical models of STH control take into account the time horizon and effectiveness of intervention required to meet WHO targets, bridging the gap between theoretical modeling and practical programming needs.

### 1.1 Rationale of the study

Although numerous mathematical models have been developed to evaluate STH control strategies, the existing modeling literature remains highly scattered. Few of the systematic reviews have addressed specific aspects—for instance, the cost-effectiveness of STH preventive chemotherapy [68]; achievement of 2020 goals for nine neglected tropical diseases using quantitative modeling [28]; the general progress of STH control strategies between 2008–2018 [39]; and complexities STH-related morbidity [7]. However, there is a notable lack of comprehensive systematic synthesis that classify mathematical models based on structural assumptions and population features, explore intervention effectiveness in accelerating the achievement of WHO targets, and predict the timeline required for an intervention to achieve elimination of STH or control their morbidity.

The shortage of an in-depth overview of the existing mathematical models for STH management is one important knowledge gap. Without a clear understanding of the types of models used, their effectiveness in reaching the WHO 2011–2020 and 2021–2030 targets, and the timelines required to achieve these goals, researchers and policymakers risk relying on models that may be inappropriate, overly simplistic, or inadequately validated for guiding programmatic decisions. This could lead to misaligned intervention strategies, inefficient resource allocation, and ultimately delays in achieving global control or elimination aims.

Consequently, the study seeks to fill identified gaps by systematically categorizing model types according to their population stratification and structural assumptions, analyzing their predictive efficacy and impact in reaching WHO targets, and determining the time required for a modeled intervention to achieve either morbidity control or STH elimination. More precisely, the study will be helpful for determining the most effective intervention to reach the 2030 eradication target.

The primary objective of the current study is guided by the following three research questions:

i. What types of mathematical models have been developed to simulate the transmission dynamics and control of soil-transmitted helminthiasis, and how do they differ in terms of structure and population features?
ii. How effective are various control strategies in terms of impact and efficiency in achieving WHO targets?
iii. What are the predicted timelines required for a modeled intervention to achieve WHO 2011–2020 and 2021–2030 NTD targets particularly STH?

## 2 Material and methods

The study employed the standard procedure for conducting a systematic review in accordance with the 2020 statement of the Preferred Reporting Items for Systematic Reviews and Meta-Analyses (PRISMA) criteria [55, 56]. The use of PRISMA guidelines is meant to lessen bias throughout the entire systematic review process and enhance the validity of the findings related to mathematical modeling of helminthiasis control strategy of the review at hand. A protocol was developed for this investigation and submitted to the PROSPERO international prospective register of systematic reviews with registration data PROSPERO 2025 (CRD420251090375). The completed PRISMA 2020 checklist and abstract are provided as supporting information (see S1 PRISMA Checklist).

### 2.1 Information sources and search terms

A comprehensive literature search was conducted between January 2015 to December 2024, where guided by PRISMA guidelines of the peer-reviewed literature in three major bibliographic databases - Web of Science, Embase, and Scopus. These databases are characterized by structured searches containing many filters, including but not limited to document type, subject area, publication type, and publication stage, ensuring an optimized search process. The databases cover a wide range of disciplines to allow capturing high-quality and peer-reviewed articles. Furthermore, additional literature was obtained through reference searches of other systematic reviews and other manuscripts. The development of the search strategy was guided by the research questions and focused on three core conceptual domains: model term, intervention and disease term as three primary key concepts employed to get the search term. Boolean operators were utilized to combine the three primary keywords across these domains, resulting in a standardized search string of the form: (“mathematical model*”) + (“soil-transmitted helminth*”) + (“control strateg*”), where the asterisk (*) serves as a truncation or wildcard operator to capture possible word variation or different ending with the given word root and and ensure comprehensive retrieval of related terms.

Further, we used alternative words, derivatives, and synonyms for each primary keyword mentioned above to expand and enhance the efficacy of the search query in retrieving the intended research papers. The complete search terms can be found in Table 1, but syntax modifications are to be made to fit the requirements of specific database.

**Table 1:** Search query components.

| Component | search terms |
| --- | --- |
| **Modeling terms** | ("mathematical model*" OR "computational model*" OR "stochastic model*" OR "quantitative model*" OR "epidem* model*" OR "theoretical model*" OR "predictive model*" OR "deterministic model*" OR "individual-based model*" OR "agent-based model*" OR "network model*" OR "network-based model*" OR "compartmental model*" OR "population model*" OR "hybrid model*" OR "computational approach*") |
| **AND** |  |
| **Control terms** | ("control strateg*" OR intervention* OR "mass drug administration" OR "preventive chemotherapy" OR sanitation OR "hygiene promotion" OR "deworming" OR "public health intervention*" OR "disease control program*" OR "health education" OR approach* OR "framework" OR program* OR guideline* OR "anthelmintic treatment*" OR "preventive intervention*" OR "helminth control") |
| **AND** |  |
| **Disease terms** | ("helminthiasis" OR "Ascari*" OR "roundworm" OR "necator americanus" OR "ancylostoma duodenale" OR "hookworm*" OR "Trichur*" OR "soil-transmitted helminth*" OR "STH" OR "helminth infection*" OR "intestinal parasite*" OR "intestinal helminth*" OR "intestinal nematode*" OR "geohelminth*" OR "whipworm" OR "Strongyloides stercoralis") |

### 2.2 Eligibility criteria

The search query was limited to only return papers published between 2015 and 2024. The limitation was put in place to concentrate on studies that used mathematical models to meet the threshold requirement for STH morbidity control or interrupting transmission. This ten-year window was selected to ensure relevance to ongoing global health efforts and to capture modeling studies that align with or contribute to the targets outlined in the WHO roadmaps, particularly those from 2011–2020 and 2021–2030. Although the selected timeframe may not encompass the full breadth of earlier literature supporting the 2021–2030 roadmap, it offers a balance between recency, methodological advancement, and practical manageability of the review scope. This recent period also enhances the quality of the review by emphasizing studies that incorporate modern modeling techniques, reflect updated epidemiological data, and are more likely to inform current policy and implementation strategies. The study employed five inclusion criteria against every research article identified from the mentioned three databases.

The inclusion criteria used in this study were as follows:

a. Any representation of parasite-host dynamics to STH transmission.
b. A compartmental or stochastic based mathematical or hybrid modeling frameworks that uses the differential equations.
c. A quantitative modeling evaluating STH control.
d. Peer-review articles.
e. Source is in English.

The exclusion criteria used were:

a. All quantitative studies that do not evaluate the control strategy
b. All qualitative models, like descriptive statistical models, which account only for probability distributions, example regression and clustering analysis.
c. All within-pathogen/host models and other studies modeling different stages of the parasite life cycle inside its host.
d. Non-quantitative models and unpublished articles.
e. Grey literature (students’ dissertations, reports, posters, conference presentations/proceedings, abstracts, letters, books, and book sections).

### 2.3 Study selection and consistency check

Initially, all research articles retrieved from the three selected databases were imported into EndNote to remove duplicates and manage references. The articles were then transferred to the Systematic Review-Accelerator tool for further duplicate removal. Addition of relevant studies were identified via manual reference list of systematic reviews and included articles. Three primary steps were involved in the inclusion of papers in this systematic review: identification of the articles, screening, and eligibility evaluation. Identified articles then proceeded to the second stage of screening. The screening process was conducted in two rounds by five reviewers. In the first round, all article titles and abstracts were assessed to ascertain their relevance. Articles were taken to the next stage if the title or abstract contained a combination of keywords related to the search terms; otherwise, they were excluded. A comprehensive evaluation of articles was done in the second round. All reviewers read the full texts of articles in relation to eligibility criteria, ready for either inclusion or exclusion from the review. Throughout the entire screening process, discrepancies were resolved through team discussion and consensus, based on the inclusion and exclusion criteria. The entire selection process and included studies can be observed in the PRISMA flow diagram (see Fig. 1).

**Fig. 1:**
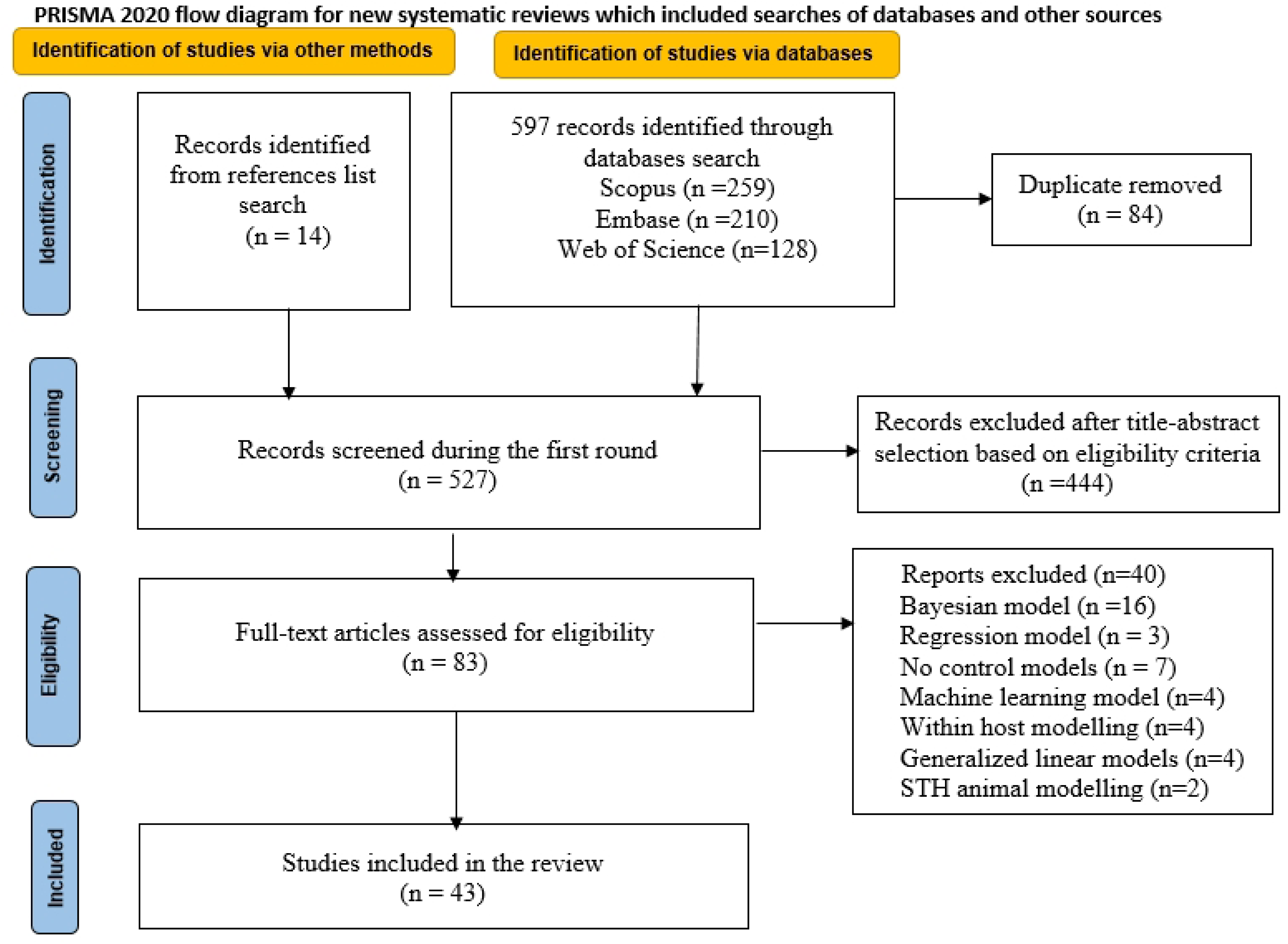
PRISMA schematic diagram.

### 2.4 Data extraction and synthesis approach

Microsoft Excel was employed for data extraction and management. The structured spreadsheet included columns capturing key elements such as study identification/characteristics, model type, intervention type, study objectives, time-bound prediction, WHO targets, predictive outcomes, sensitivity/uncertainty, effectiveness of the interventions, key findings, limitation, future work (see table 1, 2, 3, and 4). In addition, VOSviewer (version 1.6.20) was used to generate bibliometric maps and visualize co-occurrence patterns among terms and authors [69]. Qualitative synthesis was conducted by grouping mathematical models basing common design features and comparing. Also, control strategy scenarios were compared by looking their effectiveness to achieve WHO programmatic goals. In addition, either present or absent of timeframes for models were recorded and critically examined. A formal meta-analysis was not conducted due to substantial heterogeneity in modeling frameworks, intervention types, outcomes, and epidemiological assumptions across studies. Instead, we employed a narrative approach to identify recurring patterns, assess intervention impact, and highlight modeling gaps.

### 2.5 Risk of bias and assessment of study quality

The risk of bias assessment was conducted by evaluating the validity of individual studies in order to understand the implication of this risk of bias on the overall quality of evidence for a given outcome related to our study question. This study assessed the risk of bias and quality of studies using the Assessment of Modeling Studies (AMS) tool. The tool commonly used to assess methodological quality of modeling research in the fields of health and economics. We adopted and modified the tool by adding two more criteria (alignment with WHO targets and predictive timeline) to the existing criteria in the AMS [23] to enhance its relevance to the study at hand. The modification aimed to assesses whether existing mathematical models captured model’s alignments WHO targets and time-bound prediction. Models aligning to WHO targets improve their utility and specifying a predictive timeline enables decision-makers to understand the expected timeframe within which an intervention may achieve outcomes such as morbidity control or transmission elimination.

The modified AMS tool utilized the quality of study assessment format as described in [27]. All criteria utilized as per the AMS tool described the characteristics of individual research articles chosen from the review (see table 3). Number assignments from 0 to 2 were used, in which 0 represents absent, 1 indicates partially present, and 2 indicates fully present. The total points for each modeling study is 24. We calculated the percentage for each individual article and classified them into four subcategories [44]: low (*x ≤* 50%), medium (50 *< x ≤* 65%), high (65 *< x ≤* 80%), and very high (*x >* 80%). All reviewers conducted assessments independently, but where contradictions arose, discussions were held to reach consensus.

## 3 Results

### 3.1 Trends in the number of published modeling studies

Fig. 1 presents a PRISMA diagrams for the whole screening process, where a total of 611 articles were identified through database searches and manual reference list searches. A total of 84 duplicates were identified by using both EndNote and the Systematic Review Accelerator tool. At first, 66 duplicates were removed using EndNote. Further, a total of 546 deduplicated documents from EndNote were imported to the Systematic Review Accelerator, where 18 duplicates were observed and subsequently removed. 527 records were screened during the first round, where a total of 444 records were excluded after reading their titles and abstracts. Excluded articles did not provide information related to the governing inclusion criteria and scope of the review. In total, 83 research documents were earmarked for the second round of comprehensive full-text evaluation. Upon thoroughly reviewing all research articles, 40 documents were excluded for not describing deterministic, stochastic, or hybrid models, disease transmission dynamics, or evaluating STH control strategies. Finally, 43 studies met the inclusion criteria and were included in this review (see Fig. 1). We described the characteristics of all research articles and focused on the disease threshold conditions for elimination, morbidity control, intervention impact, and effectiveness as modeled in different studies.

### 3.2 Study characteristics

This overview analyzed mathematical modeling studies between 2015 and 2024 years, showing distinct features in publication output as per Fig. 2. In the early years (2015–2017), the number of publications was relatively stable, with four (9%) research articles published annually within the given range of years, while in 2018 the number of articles published rose to six (14%). However, the field of STH mathematical modeling experienced a decline in publications to only four (9%) articles for two consecutive years between 2019 and 2020. Additionally, the field experienced another increase in articles to seven (16%) in 2021 before noticing another decline to two (5%) articles in 2022. There was no identified published article in 2023 based on the applied search strategy. The year 2024 experienced the highest number of publications (8 studies, 19%). Two years (2021 and 2024) highlighted the peaks in publication (7 and 8 articles respectively), while one year, 2022, experienced a low turn in publication output (only 2 publications). Generally, the publication output indicates fluctuating trends in publication efforts over the decade. Two years (2021 and 2024) experienced the highest number of publications, indicating heightened research work during this period, while only 2 studies were published in 2022, which may reflect delays in research output due to the COVID-19 pandemic or shifts in funding priorities. [50]

**Fig. 2:**
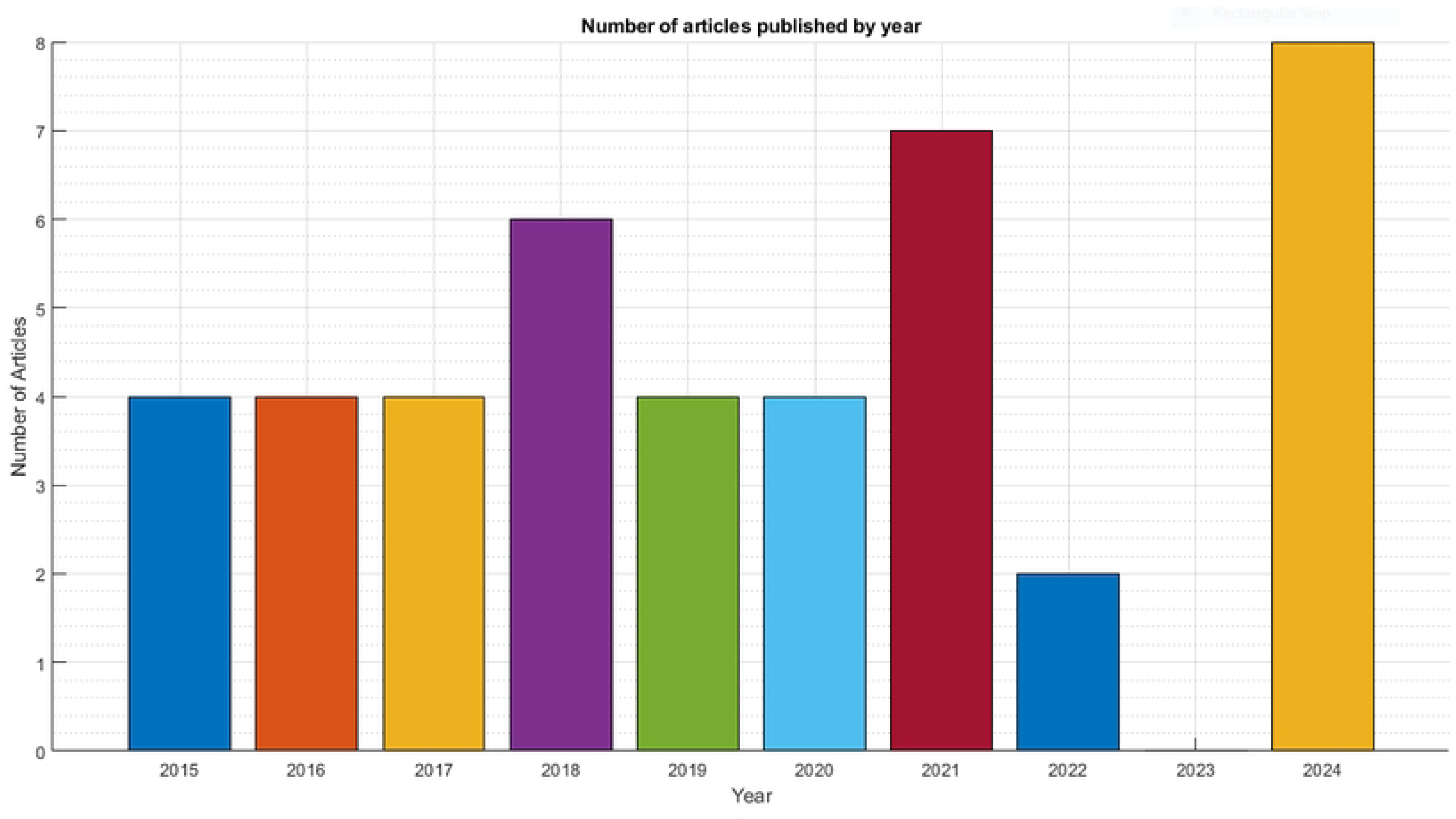
Research publication output yearly.

### 3.3 Model types by structure

In gaining a meaningful and comparing developed mathematical models for transmission dynamics and control of STH, it is crucial to explore their structures and assumptions. Structural assumptions enabled the 43 reviewed studies to be categorized into deterministic, stochastic, and hybrid models, as observed in Fig. 3. Further, table 2 entails the details on model classification based their structural assumptions.

**Fig. 3:**
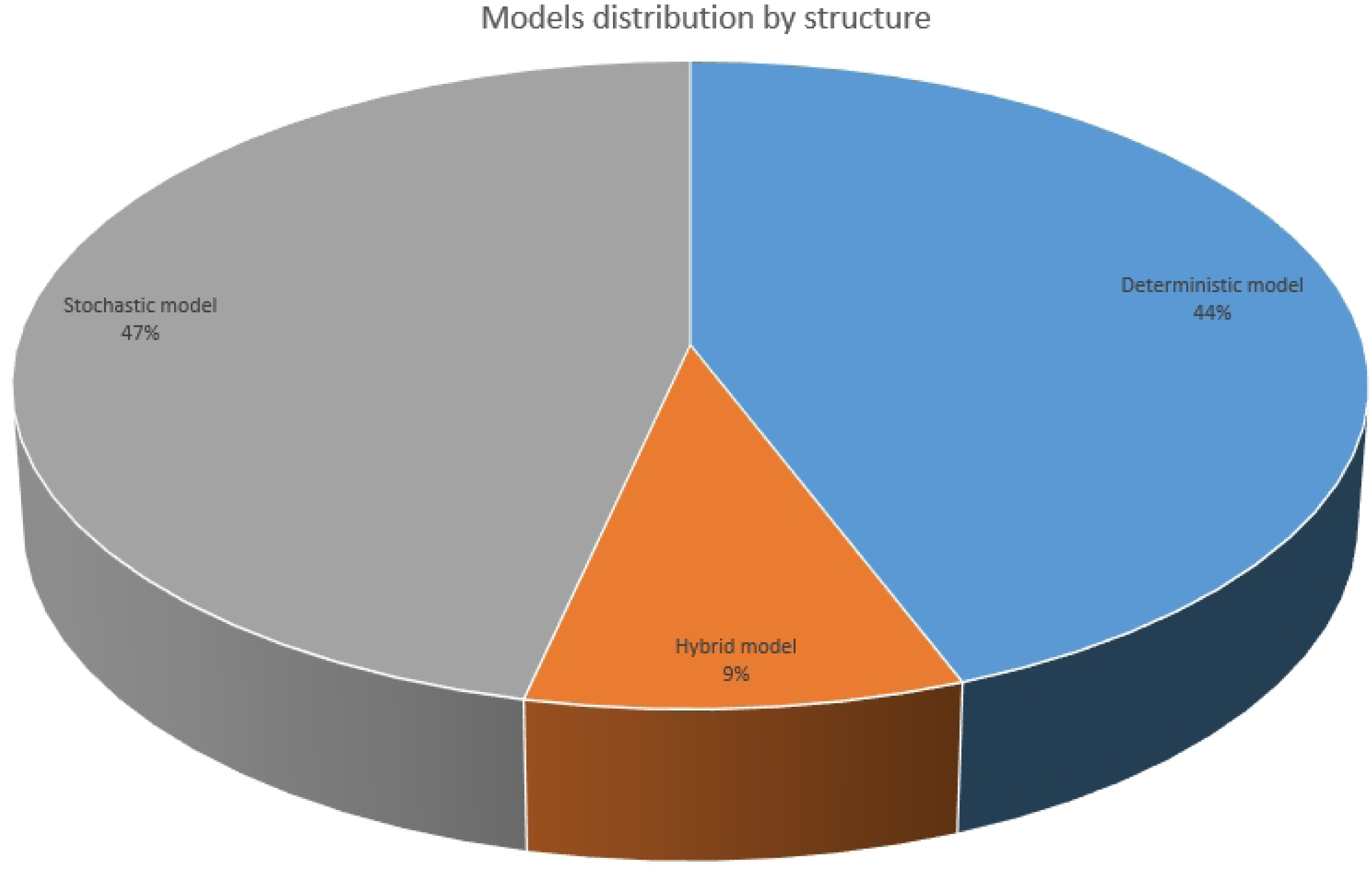
Modeling category per structure.

**Table 2:** Summary of included studies by objective, model type with design and population features, intervention type, and timeline of modeled intervention. Abbreviations: (DM = deterministic model, SD = stochastic model, SBD = school-based deworming, CBD = community-based deworming, PC = preventive chemotherapy, WASH = water sanitation and hygiene)

| Objectives | Author | Year | Model Type | Intervention type | Structured population model | Time line (years) |
| --- | --- | --- | --- | --- | --- | --- |
| Predicting impact of targeted and mass deworming on STH transmission using Worm-isim | Clarke et al. [11] | 2019 | SD | SBD & CBD | Age structured | - |
| Validation of ICL & EML models for STH by comparing their predictions | Coffeng et al. [15] | 2017 | Hybrid | SBD & CBD | Age structured | - |

Table 2. cont
| Objectives | Author | Year | Model Type | Intervention type | Structured population model | Time line (years) |
| --- | --- | --- | --- | --- | --- | --- |
| Investigate trial stopping criteria in a situation where interruption of transmission is unlikely | Werkman et al. [76] | 2018 | SD | SBD & CBD | Age structured | - |
| Explore the impact of targeted deworming | Vegvari et al. [71] | 2021 | SD | SBD & WRA | Age structured | by 2030 |
| Application of fractional operator to investigate the dynamics of Hookworm infection | Mustapha et al. [41] | 2020 | DM | PC & WASH | Flat | - |
| Assess the impact of systematic non-compliance MDA programs for controlling STH | Farrell and Anderson [21] | 2018 | SD | PC | Age structured | - |
| Investigate the impact of COVID-19-related interruptions on STH control programs | Malizia et al. [35] | 2020 | SD | SBD & CBD | Age structure model | by 2030 |
| Investigate the impact of current WHO guidelines on achieving STH morbidity goals in SAC | Farrell et al. [22] | 2018 | SD | SBD & CBD | Age structured | 5-6 |
| Review how uncertainty in key assumptions on STH population biology can affect the planning and evaluation of STH control programs | Vegvari et al. [72] | 2021 | SD | SBD & WRA | Spatial | - |
| Investigate hygiene-conscious compartment | Oguntolu et al. [45] | 2023 | DM | PC & WASH | Flat | - |

Table 2. cont
| Objectives | Author | Year | Model Type | Intervention type | Structured population model | Time line (years) |
| --- | --- | --- | --- | --- | --- | --- |
| Analyze the effectiveness of different control strategies in reducing helminthiasis | Lambura et al. [31] | 2020 | DM | PC & WASH | Flat | - |
| Propose STH-Mtb co-dynamic model and explore their interaction | Lambura et al. [32] | 2020 | DM | PC | Flat | - |
| Investigate the dynamics of hookworm infection | Chazuka et al. [8] | 2024 | DM | PC & WASH | Flat | - |
| Develop and analyze an optimal control model to determine the most effective strategy for STH elimination | Okoyo et al. [48] | 2022 | DM | PC & WASH | Age structured | - |
| Evaluate the economic feasibility of expanding hookworm control strategies to community | Turner et al. [66] | 2015 | DM | SBD & CBD | Age structured | 15 |
| Develop a framework for describing the dynamics of STH transmission near the break-point | Hardwick et al. [25] | 2020 | Hybrid | PC & WASH | Flat | - |
| Develop mathematical model and investigate the factors for evolution of anthelmintic resistance | Patel et al. [57] | 2024 | DM | PC | Flat | - |
| Analyze the transmission dynamics of hookworm infection and estimate the $R_0$ | Truscott et al. [63] | 2019 | DM | SBD & CBD | Spatial | - |
| Determine the optimal control strategy for STH elimination | Chong et al. [10] | 2021 | DM | SBD & CBD | Flat | - |

Table 2. cont
| Objectives | Author | Year | Model Type | Intervention type | Structured population model | Time line (years) |
| --- | --- | --- | --- | --- | --- | --- |
| Investigate the spread of hook-worm disease and assess chemotherapy effectiveness | Kasia and Shengqiang [29] | 2016 | DM | Dual therapy | Flat | 5 |
| Explore the impact of human population movement on STH elimination efforts | Vegvari et al. [73] | 2019 | SD | SBD & CBD | Spatial | - |
| Estimate the cost-effectiveness of MDA with ivermectin for strongyloidiasis control | Coffeng et al. [13] | 2024 | SD | Dual therapy | Age structured | 5-6 |
| Identify optimal threshold statistics for the elimination of hookworm | Truscott et al. [64] | 2017 | SD | SBD & CBD | Both structure | - |
| Predict the feasibility of achieving $\leq 1\%$ prevalence of M&H hookworm infection by 2020 | Coffeng et al. [12] | 2015 | SD | PC & WASH | age structured | 2 |
| Assess the feasibility of interrupting the transmission of STH through MDA | Truscott et al. [62] | 2021 | SD | PC | Spatial | 10 |
| Assess cost-effectiveness of scaling up SBD for *Ascaris lumbricoides* by a dynamic model. | Turner et al. [67] | 2016 | SD | SBD | Age structured | 35 |
| Evaluate the impact of WASH interventions on STH transmission | Coffeng et al. [16] | 2018 | SD | PC & WASH | Age structured | - |

Table 2. cont
| Objectives | Author | Year | Model Type | Intervention type | Structured population model | Time line (years) |
| --- | --- | --- | --- | --- | --- | --- |
| Investigate the risk and speed of drug resistance in human STH populations | Coffeng et al. [14] | 2024 | SD | PC | age structured | - |
| Compare the predictions of IBM with their deterministic counterparts | Truscott et al. [61] | 2016 | Hybrid | PC | Age structured | - |
| Investigate how well standard DM match IBM simulations | Hardwick et al. [26] | 2021 | SD | PC | Spatial | - |
| Review the epidemiology of Strongyloidiasis and simulate the impact of repeated MDA | Collyer and Anderson [17] | 2023 | SD | PC | Age structured | - |
| Examine the dynamics of the parasite infection model with their stability | Chong et al. [9] | 2022 | Hybrid | PC & WASH | Flat | - |
| Analyze the compliance data of the Geshiyaro project and determine its impact on elimination | Maddren et al. [34] | 2024 | SD | SBD & CBD | Age structured | by 2028 |
| Investigate the impact of achieving WHO's targets with SBD | Truscott et al. [60] | 2015 | DM | PC | Age structured | by 2020 |
| Investigate the impact of LF MDA treatment on the prevalence of hookworm infection | Werkman et al. [75] | 2017 | DM | PC | Age structured | - |
| Predict the mean worm burden levels in different age groups over time | Okoyo et al. [47] | 2021 | DM | PC & WASH | Age structured | 5 |

Table 2. cont
| Objectives | Author | Year | Model Type | Intervention type | Structured population model | Time line (years) |
| --- | --- | --- | --- | --- | --- | --- |
| Extending classical integer-order model into fractional order model | Omede et al. [51] | 2024 | DM | WASH | Flat | - |
| Evaluate the impact for co-administration of ivermectin | Turner et al. [65] | 2016 | DM | Dual therapy | Age structured | 2 |
| Quantify how diagnostic sensitivity affects the optimal prevalence threshold for declaring interruption | Werkman et al. [74] | 2018 | SD | PC | Age structured | - |
| Propose a novel model for *A. lumbricoides* transmission with seasonal elements | Davis et al. [18] | 2018 | DM | PC | Flat | - |
| Assess the predictive performance of various sampling designs for evaluating the morbidity target | Giardina et al. [24] | 2019 | SD | PC | Spatial | - |
| Assessing different design of cluster randomized trials for MDA for STH elimination | Anderson et al. [4] | 2017 | SD | SBD & CBD | Spatial | 5 |
| Should the treatment target on community wide elimination rather than morbidity control for children? | Anderson et al. [5] | 2015 | DM | SBD & CBD | age structured | 7 |

**Table 3:** Description of the quality assessment of the included studies adopting the tool Assessment for Modelling Studies (ASM). The values represent the AMS criteria: 0 = absent, 1 = partially present, 2 = fully present.

| Author | Clear aims and objective | Intervention comparators | Outcome measure defined | Model Structure & flow chart | WHO targets alignment | Parameters Specified | Assumptions Justification | Data quality & Sensitivity/Uncertainty | Model Validation | Results Presentation | Results Discussion & Interpretation | Predictive timeline to WHO targets | Total score | Score (%) | Quality category |
| --- | --- | --- | --- | --- | --- | --- | --- | --- | --- | --- | --- | --- | --- | --- | --- |
| Clarke et al. [11] | 2 | 2 | 2 | 0 | 2 | 2 | 2 | 2 | 1 | 2 | 2 | 0 | 19 | 79.2 | High |
| Coffeng et al. [15] | 2 | 2 | 2 | 0 | 2 | 1 | 2 | 1 | 2 | 2 | 2 | 0 | 18 | 75 | High |
| Werkman et al. [74] | 2 | 2 | 2 | 1 | 2 | 2 | 2 | 1 | 2 | 2 | 2 | 0 | 20 | 83.3 | Very high |
| Vegvari et al. [71] | 2 | 2 | 2 | 0 | 2 | 2 | 2 | 2 | 1 | 2 | 2 | 0 | 19 | 79.2 | High |
| Mustapha et al. [41] | 2 | 2 | 2 | 2 | 2 | 2 | 1 | 2 | 1 | 2 | 2 | 0 | 21 | 87.5 | Very high |
| Farrell and Anderson [21] | 2 | 1 | 2 | 1 | 2 | 1 | 2 | 2 | 1 | 1 | 2 | 0 | 19 | 79.2 | High |
| Malizia et al. [35] | 2 | 2 | 2 | 1 | 2 | 2 | 2 | 2 | 1 | 2 | 2 | 2 | 22 | 91.6 | Very high |
| Farrell et al. [22] | 2 | 2 | 2 | 1 | 2 | 2 | 2 | 2 | 2 | 2 | 2 | 2 | 22 | 91.6 | Very high |
| Vegvari et al. [72] | 2 | 2 | 2 | 1 | 2 | 1 | 2 | 1 | 1 | 2 | 2 | 2 | 20 | 83.3 | Very high |
| Oguntolu et al. [45] | 2 | 1 | 2 | 2 | 2 | 2 | 1 | 1 | 0 | 2 | 2 | 0 | 17 | 70.8 | High |
| Lambura et al. [32] | 2 | 2 | 2 | 2 | 2 | 1 | 2 | 1 | 0 | 2 | 2 | 0 | 17 | 70.8 | High |
| Lambura et al. [31] | 2 | 2 | 2 | 2 | 2 | 2 | 2 | 1 | 0 | 2 | 2 | 0 | 19 | 79.2 | High |
| Chazuka et al. [8] | 2 | 2 | 2 | 2 | 2 | 2 | 2 | 1 | 0 | 2 | 2 | 0 | 19 | 79.2 | High |

**Table 3 – continued from previous page**
| Author | Clear aims and objective | Intervention comparators | Outcome measure defined | Model Structure & flow chart | WHO targets alignment | Parameters Specified | Assumptions Justification | Data quality & Sensitivity/Uncertainty | Model Validation | Results Presentation | Results Discussion & Interpretation | Predictive timeline to WHO targets | Total point | Total score (%) | Quality category |
| --- | --- | --- | --- | --- | --- | --- | --- | --- | --- | --- | --- | --- | --- | --- | --- |
| Okoyo et al. [48] | 2 | 2 | 2 | 1 | 2 | 2 | 2 | 2 | 1 | 2 | 2 | 0 | 20 | 83.3 | Very high |
| Turner et al. [66] | 2 | 2 | 2 | 1 | 2 | 1 | 2 | 2 | 1 | 2 | 2 | 0 | 19 | 79.2 | High |
| Hardwick et al. [25] | 2 | 2 | 2 | 1 | 2 | 2 | 1 | 1 | 1 | 2 | 2 | 0 | 18 | 75 | High |
| Patel et al. [57] | 2 | 1 | 2 | 1 | 2 | 2 | 1 | 0 | 0 | 2 | 2 | 0 | 15 | 62.5 | Medium |
| Truscott et al. [63] | 2 | 2 | 2 | 1 | 2 | 2 | 1 | 2 | 1 | 2 | 2 | 0 | 19 | 79.2 | High |
| Chong et al. [10] | 2 | 1 | 2 | 1 | 2 | 1 | 2 | 1 | 1 | 2 | 2 | 0 | 17 | 70.8 | High |
| Kasia and Shengqiang [29] | 2 | 2 | 2 | 2 | 2 | 1 | 2 | 2 | 2 | 2 | 2 | 2 | 23 | 95.8 | Very high |
| Vegvari et al. [73] | 2 | 2 | 2 | 1 | 2 | 1 | 2 | 1 | 1 | 2 | 2 | 0 | 18 | 75 | High |
| Coffeng et al. [14] | 2 | 2 | 2 | 1 | 2 | 2 | 1 | 2 | 1 | 2 | 2 | 2 | 21 | 87.5 | Very high |
| Truscott et al. [64] | 2 | 2 | 2 | 1 | 2 | 1 | 2 | 2 | 1 | 2 | 2 | 0 | 19 | 79.2 | High |
| Coffeng et al. [12] | 2 | 2 | 2 | 1 | 2 | 2 | 1 | 2 | 2 | 2 | 2 | 2 | 23 | 95.8 | Very high |
| Truscott et al. [62] | 2 | 1 | 2 | 1 | 2 | 2 | 2 | 2 | 2 | 2 | 2 | 2 | 23 | 95.8 | Very high |
| Turner et al. [65] | 2 | 1 | 2 | 1 | 2 | 2 | 2 | 2 | 1 | 2 | 2 | 2 | 21 | 87.5 | Very high |
| Coffeng et al. [16] | 2 | 2 | 2 | 1 | 2 | 1 | 2 | 2 | 1 | 2 | 2 | 0 | 19 | 79.2 | High |
| Coffeng et al. [13] | 2 | 2 | 2 | 2 | 2 | 1 | 2 | 1 | 1 | 2 | 2 | 0 | 19 | 79.2 | High |
| Truscott et al. [61] | 2 | 1 | 2 | 1 | 2 | 2 | 2 | 2 | 1 | 1 | 2 | 0 | 18 | 75 | High |
| Hardwick et al. [26] | 2 | 1 | 2 | 1 | 2 | 2 | 2 | 1 | 1 | 2 | 2 | 0 | 18 | 75 | High |
| Collyer and Anderson [17] | 2 | 1 | 2 | 2 | 2 | 1 | 1 | 2 | 1 | 2 | 2 | 0 | 18 | 75 | High |
| Chong et al. [9] | 2 | 2 | 2 | 1 | 2 | 2 | 1 | 2 | 1 | 2 | 2 | 0 | 19 | 79.2 | High |
| Maddren et al. [34] | 2 | 2 | 2 | 0 | 2 | 2 | 1 | 2 | 2 | 1 | 2 | 2 | 19 | 79.2 | High |
| Truscott et al. [60] | 2 | 2 | 2 | 1 | 2 | 1 | 2 | 2 | 2 | 2 | 2 | 2 | 23 | 95.8 | Very high |
| Werkman et al. [75] | 2 | 2 | 2 | 1 | 2 | 2 | 1 | 1 | 1 | 2 | 2 | 0 | 18 | 75 | High |
| Okoyo et al. [47] | 2 | 2 | 2 | 2 | 1 | 1 | 2 | 2 | 1 | 2 | 2 | 2 | 23 | 95.8 | Very high |

**Table 3 – continued from previous page**
| Author | Clear aims and objective | Intervention comparators | Outcome measure defined | Model Structure & flow chart | WHO targets alignment | Parameters Specified | Assumptions Justification | Data quality & Sensitivity/Uncertainty | Model Validation | Results Presentation | Results Discussion & Interpretation | Predictive timeline to WHO targets | Total point | Total score (%) | Quality category |
| --- | --- | --- | --- | --- | --- | --- | --- | --- | --- | --- | --- | --- | --- | --- | --- |
| Omede et al. [51] | 2 | 1 | 2 | 2 | 2 | 1 | 0 | 1 | 0 | 2 | 2 | 0 | 15 | 62.5 | Medium |
| Turner et al. [67] | 2 | 2 | 2 | 1 | 2 | 1 | 2 | 1 | 1 | 2 | 2 | 0 | 18 | 75 | High |
| Werkman et al. [76] | 2 | 2 | 2 | 1 | 2 | 1 | 1 | 1 | 1 | 2 | 2 | 0 | 17 | 70.8 | High |
| Davis et al. [18] | 2 | 2 | 2 | 2 | 2 | 2 | 2 | 1 | 2 | 2 | 2 | 0 | 21 | 87.5 | Very high |
| Giardina et al. [24] | 2 | 2 | 2 | 1 | 2 | 2 | 1 | 1 | 2 | 2 | 2 | 0 | 20 | 83.3 | Very high |
| Anderson et al. [4] | 2 | 2 | 2 | 0 | 2 | 2 | 1 | 2 | 1 | 2 | 2 | 2 | 20 | 83.3 | High |
| Anderson et al. [5] | 2 | 2 | 2 | 1 | 2 | 2 | 2 | 1 | 2 | 2 | 2 | 2 | 22 | 91.7 | Very high |

### Deterministic

Deterministic models were 19 out of 43 reviewed studies (44%). These are the framework employing differential equations (often ordinary differential equations) to predict the dynamics of STH under fixed model parameters and averaged population behavior. Of these, 12 research articles (63%) of deterministic models published between 2015 and 2020 with the reduced number (7 studies) appearing to be published between 2021 and 2024. These models offer several advantages, including simplicity in doing theoretical analyses such as equilibria, stability conditions, and bifurcation. In addition, these frameworks are cheap in computational simulation and require less memory. Most of the reviewed deterministic studies were typically a normal SEIR (Susceptible–Exposed–Infected–Recovered) compartment model for human population-level dynamics with the addition of other compartments having parasite information (like worm burden intensity and larva/egg stages) [31, 32, 45, 8, 18, 57, 10, 29]. Eight studies out of 19 were age-structured deterministic models [47, 48, 5, 65, 75, 60, 66, 65]. Only one study examined the heterogeneity in both age and space [63]. Two studies of the 19 were fractional order models capturing memory behavior and anomalous diffusion [51, 41] using Caputo derivative type. The memory kernel used these models based on power-law and Mittag-Leffler [41, 51], respectively. Further, few deterministic modeling studies evaluated economic modeling and effectiveness of interventions for long-term STH transmission dynamics. For instance, the researchers in [8, 31, 45] defined the strategic approach for Hookworm mitigation, assessment of *R*_0_, and stability equilibria and studied the optimal control of STH.

### Stochastic models

Stochastic models were the most commonly utilized framework (20 studies, 47%) to account for variability and randomness in many processes such as biological, environmental, and social phenomena. Nearly half (9 studies) of modeling work were published between 2020 and 2024, reflecting increasing recognition of the importance of uncertainty and heterogeneity in modeling STH transmission dynamics. Randomness in modeling enhances suitability in capturing uncertainties in transmission dynamics and intervention outcomes. Stochastic models are suited for small populations with high variability, systems driven by noise, and situations where probabilistic outcomes matter. These models are applicable in rare events such as resistance emergence due to low-probability mutations, and disease fade-out due to chance—dynamics that may be missed in deterministic model frameworks. Two stochastic models used an event-driven approach with Gillespie-type algorithms and investigated discrete transmission events [26, 74]. Another study focused on Markovian egg pulse events and migration-driven reintroduction [26]. Seven of the stochastic models were metapopulational [4, 24, 26, 62, 72, 63, 71]. Additionally, eleven of the stochastic models were individual-based models (IBM) which simulated treatment history, acquisition rate of worms, and environmental contamination at the individual level [11, 71, 21, 22, 35, 64, 16, 14, 17, 34, 76]. These IBMs improved disease understanding and predicted elimination thresholds. For example, the study in [74] conducted research defining the stopping criteria for ending randomized clinical trials.

### Hybrid

Hybrid models are cohesive frameworks with both deterministic and stochastic characteristics to capture disease transmission dynamics across multi-scale. The multi-scale modeling enhance a comprehensive capturing of disease dynamics heterogeneity. Both deterministic and stochastic components of a hybrid used to capture different behavior, for example STH mean worm burden and prevalence can be studied through deterministic component, while individual variation in treatment and worm acquisition can be explored via a stochastic component is needed [61]. The model was classified hybrid for having either both stochastic and deterministic components or by having a frameworks that compare results coming from both components.

There were only four (9%) studies identified to employ hybrid frameworks [25, 61, 9, 15]. Of these, three studies incorporated stochastic perturbation into their primary deterministic models [25, 61, 9], while only one study had a separated stochastic part with deterministic but compared their outcomes [15]. Since the intention of classification is to see the outcome when the methodology differs, this reason led us to consider the mentioned study as a hybrid model. Further, elimination feasibility has been evaluated using these models, where baseline prevalence was captured deterministically while movement was captured by the stochastic component [25]. Notably, there are only two studies that explicitly modeled human movement as a stochastic entity while the main transmission dynamics were captured by the deterministic component [9, 25]. Additionally, one hybrid model sampled the parameters stochastically and modeled prevalence of transmission deterministically [63]. These modeling studies considered contamination and reservoir dynamics to be deterministic, and the host treatment history and WASH adherence to be stochastic.

### Modeling classification by population stratification

The current study further classified the reviewed studies depending on population representation. This key aspect is critical in understanding whether the reviewed models treated the population as homogeneous or incorporated some elements of heterogeneity in age or space (spatial distribution). The overview at hand classified the models based on population stratification into three categories: non-structured models, age-structured models, and spatial models (see Fig. 4).

**Fig. 4:**
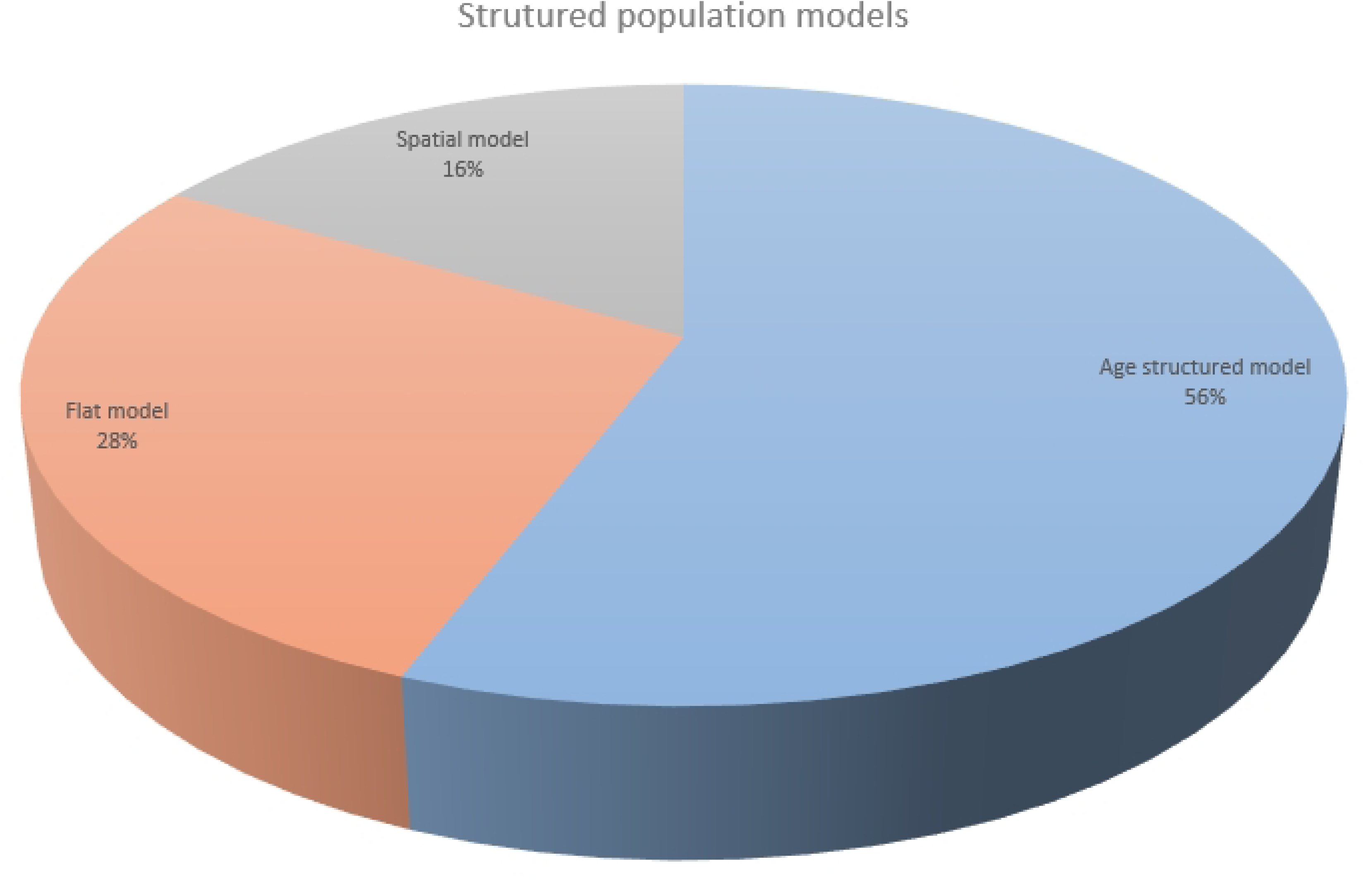
Models category by population presentation.

### Flat structured models

These are models which do not consider internal stratification of the population. They treat individuals in every compartment as having the same biological, demographic, and behavioral properties. Non-structured frameworks do not account for differences in age, behavior, or immunity. This simplification allows for analytical tractability but may limit the model’s applicability to age-targeted control programs.

The studies categorized as flat structured models (12 articles, 28%) were mostly deterministic frameworks (refers to table 2). Of these, 10 flat frameworks were of deterministic while 2 were hybrid. Additionally, only a single out of 12 studies explored co-infection particularly STH-TB co-interaction [32]. Similarly, one other study investigated the seasonal impact (temperature and rainfall) on timing of MDA for STH [18]. The majority of the unstructured models acknowledged the fact of heterogeneity as a limitation that needs to be addressed.

### Age structured models

Age-structured models (24 studies, 56%) are frameworks which divide the population into age groups. These models divide the population into either discrete or continuous age cohorts to capture differences in treatment rate, exposure, infection rate and drug efficacy across age groups. Of these age structured models, 14 were stochastic frameworks, 8 were deterministic and 2 hybrid age structured models. These models consider parameters such as contact rate and treatment coverage to vary by age. The reviewed studies of age-structured models divided the population into either four or three age cohorts. Notably, two studies considered 4 age strata: infants (0–1 years), PSAC (2–4 years), SAC (5–14 years), and adults (older than 15 years) [66, 67]. Biological processes such as exposure contact rate peak at the age of 5–15 years for *A. lumbricoides* but maintain linearity in adulthood for hookworm [60] but *T. trichiura* shows a bimodal pattern for 5–15 years and 30–50 years [24]. The study in [64] demonstrated that worm acquisition from the infectious material is mediated by age-dependent contact rate and leads to the age profile of infection for parasitic worms.

After population stratification, age-specific exposure profiles were explored using either ordinary differential equations or partial differential equations. Partial differential equations consider age bins as a continuous variable, while ordinary differential systems treat them as discrete variables. These modeling approaches facilitated the determination of age-specific transmission patterns and evaluate the differential MDA impact across age groups. For instance, the study revealed that school-based deworming is effective against *A. lumbricoides* and that 10 years is required for elimination [60]. Age-structured models improved understanding of species-specific dynamics, STH elimination feasibility, and informed combination therapy. For instance, one study reported that child treatment strategy is effective for *A. lumbricoides* [60] and elimination is feasible with 75% SAC coverage [65].

### Spatial models

Spatial models consider a population structure and heterogeneity of the STH in a space. They inform on how STH evolves in space and time. There are six (7 articles, 16%) spatial frameworks among the 43 reviewed articles [4, 72, 63, 73, 62, 26, 24]. Of these, 6 studies were spatial stochastic models and one was spatial deterministic. These models capture geographical variation in local transmission, migration, and environmental factors. For instance, a study in [73] explored the impact of seasonal human movement within a two-village metapopulation with an intention to eliminate STH and reported that even a small migration rate (2%) from high-prevalence to low-prevalence areas affected elimination probability by 50%. Also, researchers in [25] demonstrated that migration rates exceeding the turnover of the infectious reservoir undermine the local preventive chemotherapy effort, leading to rebound risks.

The probability of disease elimination depends on cluster size and villages. For example, one study demonstrated that elimination probability is nearly zero when the cluster size is 2000 individuals [73, 62]. Additionally, An increased number of villages in a cluster undermines the chance of transmission interruption at the cluster level [75]. The disease dynamics are studied under small village units, excluding large villages, since small villages form good epidemiological clusters [74]. Movement between locations to account for local populations can be a good approach to study elimination prospects.

### 3.4 Interventions types

Over many years, a variety of control strategies have been incorporated in mathematical modeling studies to explore their impact and effectiveness on prevalence reduction and elimination feasibility of STH. Preventive chemotherapy (PC), integration of PC and WASH, targeted deworming (for school adolescence girls and women of reproductive age (WRA)), combination therapy, school-based deworming (SBD), and community-based deworming (CBD) were the categories of interventions employed in 43 reviewed studies (see Fig. 5 and table 2).

**Fig. 5:**
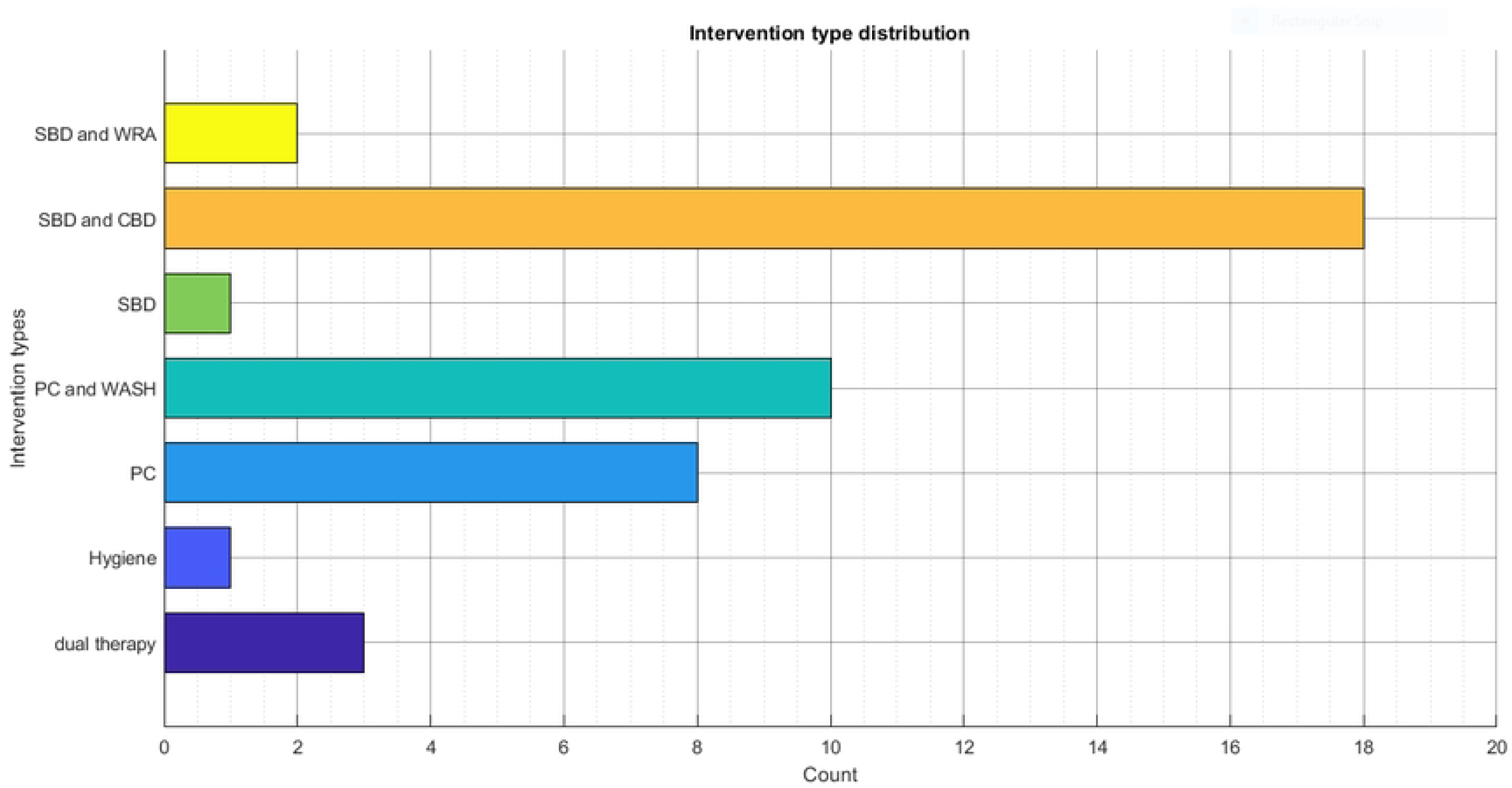
Intervention types.

Out of the 43 studies, 18 focused on comparing school-based deworming and community-wide MDA against STH infections [5, 4, 24, 76, 75, 60, 34, 14, 64, 73, 63, 66, 72, 22, 35, 74, 15, 11], while only one study employed school-based MDA [67]. Similarly, eight modeling studies employed preventive chemotherapy (PC) programs to curtail STH [21, 57, 10, 62, 61, 17, 18, 26]. There are only three studies on dual therapy [65, 13, 29]. Further, two studies examined targeted deworming (SBD and WRA) [71, 12], while 10 studies modeled integrated approach with PC and WASH [41, 31, 8, 48, 25, 16, 9, 47, 51, 32]. Notably, only one study investigated hygiene consciousness in a specific compartment of a deterministic model [45].

### School based versus community wide deworming

School-based deworming (SBD) refers to preventive chemotherapy administered via mass drug administration (MDA) to children aged 2–5 years (classified as pre-school-age children, PSAC) and school-aged children (SAC: 5–15 years) [47, 66, 22]. This intervention was administered either annually or biannually, depending on the prevalence level in the specific endemic setting, as per WHO guidelines (annual if prevalence is 20–50% and semi-annual if higher than 50%).

The reviewed studies showed a substantial variation on the impacts and effectiveness of SBD in achieving WHO targets. For example, one study highlighted that semi-annual deworming of children in high-transmission settings (with *R*_0_ *>* 2) substantially reduced reinfection risk caused by untreated adults [76], though there is a chance of drug resistance development [14]. Moreover, another study [24] confirmed that school-based semiannual PC was highly effective for all species, nearly always meeting the morbidity target. Additionally, other modeling studies highlighted prevalence reduction to a certain threshold but reaching elimination remain as a problem. For instance, Truscott et al. [60] revealed that school-based deworming reduces the overall worm burden by 22%. Similarly, a study by [76] showed that high PSAC/SAC coverage (90%) increases the likelihood of elimination to 71.7%. Further, researchers in [11] reported that school-based deworming reduces helminthiasis morbidity but is unlikely to achieve elimination due to persistent adult infections. Treating only high-risk groups, particularly children, reduces the mean worm burden [10, 29]. Furthermore, modeling study by Vegvari et al. [73] suggested that eliminating *A. lumbricoides* is possible with interventions targeting SAC.

Community-wide mass drug administration (MDA) refers to deworming programs expanded to all age groups, including preschool-age children (2–5 years), school-aged children (5–15 years), and adults (*≥*15 years) [48, 24, 66]. This deworming program involves the delivery of preventive chemotherapy through mass drug administration across a broad community. Examined modeling studies revealed significant variation in the time required for elimination, transmission interruption, or morbidity control. For instance one study reported that communitywide preventive chemotherapy reduces prevalence to near zero within 5-6 years [13]. Another study demonstrated that targeting the wider community via MDA shortens elimination time by 2.1 years [35]. Community-wide treatment achieved elimination quickly, while child-targeted interventions failed to break transmission within the same timeframe [66]. In addition, the likelihood of eliminating STH is higher when community preventive chemotherapy is administered in small clusters rather than large ones [64].

The higher coverage and frequency improved elimination efforts of the disease even in high transmission settings. For instance, one study highlighted that 3 rounds of community MDA at *≥*75% are required for elimination prospects given *R*_0_ = 2.5, and 9 rounds with *R*_0_ = 5 [66]. Preventive chemotherapy at 90% and 80% for school and pre-school aged children and adults, respectively, interrupted the transmission of STH to less than 3% 6 months after the last round of MDA [74]. Researchers in [24] reported that community-wide PC (annual or semi-annual) consistently achieved the morbidity target, even in high-prevalence settings. The community-wide deworming is cost-effective (317-748% more than SBD) in interrupting STH transmission, but it is logistically challenging to implement [66].

### Targeted deworming

It is a kind of deworming targeted to women of reproductive age (WRA: 15-50 years) and adolescent girls (15-19 years). The studies treated adolescent girls of the mentioned age as school-based deworming. Studies reported SBD as an intervention for morbidity reduction for adolescent girls. There are two modeling studies that dealt with targeted deworming [71, 12].

One study revealed that the combination approach of SBD and WRA treatment works better than an independent control, particularly in elimination prediction [71]. Researchers in [71] demonstrated that annual deworming of adolescent girls (15-19) reduced moderate to high intensity hookworm infection by 60% in moderate settings using the Erasmus medical Center of Rotterdam (EMC) model and only 12-27% in high transmission settings by both models (EMC and Imperial College in London (ICL)). Moreover, treatment of WRA (15-50) during pregnancy or lactation (2 times in prenatal and 1 time for postnatal) reduced M&HI infections by only 20% by both models (ICL/EMC). This reduction seems small but significant for reducing anemia and improving child birth weight [71]. Coffeng et al. [12] reported that helminth infections can be controlled by preventive chemotherapy for both school children and women of childbearing age (WCBA). The study reported that including WCBA in the current control strategy improves achievement of the goals within 2 years in low and moderate settings, but in higher transmission settings, health education or WASH will facilitate goal attainment [12].

### Integrated PC with WASH

We examined articles that modeled preventive chemotherapy via MDA complemented by water, sanitation, and hygiene (WASH), footwear, health education, and other behavioral interventions. Notably, most studies (9 articles, 90%) which modeled these integration approaches were conducted beyond 2020. Only one study modeled integrated PC with WASH before 2020. A study in [48] showed that combining MDA targeted to communities with high WASH coverage (95%) is more efficient as they synergistically reduce transmission and environmental contamination. Also, researchers in [8] demonstrated in their modeling activity that shoe-wearing habits implemented with preventive chemotherapy are effective in reducing transmission intensity. The WORMSIM framework was employed to assess the effect of combining preventive chemotherapy with WASH at school and community levels [16]. The short- and long-term effects of WASH modalities implemented alongside PC were evaluated under the mentioned framework. While, the short-term impact of WASH is masked by PC due to rapid drug effects, but long-term WASH is critical to sustaining gains from PC. Notably, hookworm is about 4 times as much slower to respond to WASH compared to other helminth species. Similarly, researchers in [12] confirmed that both PC and WASH control strategies improve the probability of achieving WHO’s <1% M&HI infections goal, while a study in [41] reported that treatment using PC with sanitation is highly effective in reducing heavy infections. Moreover, health literacy for women of reproductive age leads to an additional 12% reduction in prevalence [71] while hygiene education reduces reinfection by 17% [41].

### Combination therapy

Dual-therapy strategies involves combining albendazole with either ivermectin or levamisole to improve drug efficacy against *T. trichiura* and hookworm treatment [29, 65]. Turner et al. [66] simulated the co-administration of albendazole with ivermectin and reported an egg reduction rate (ERR) of 92–95% for *T. trichiura*, compared to 60% with albendazole monotherapy. Similarly, Kasia and Shengqiang [29] predicted a 92.5% efficacy for hookworm when albendazole was combined with levamisole. Further, Turner et al. [65] highlighted in their study that expanding ivermectin-albendazole co-administration to adults could reduce the elimination timeline to two years. In addition, semiannual treatment disturbs the parasite population before eggs are introduced to the environment [18].

### Preventive chemotherapy

Several of the non-structured frameworks in this study modeled preventive chemotherapy by assuming the transmission dynamics to be uniform across all age groups. For instance, the modeling study by Davis et al. [18] reported a reduction in mean worm burden by timing preventive chemotherapy. In addition high treatment frequency with preventive chemotherapy improves outcomes in settings with systematic non-compliance [21]. Treating less frequently with PC while maintaining high coverage reduces the worm burden given time frame is long [57]. However, researchers in [57] reported that repeated rounds of treatment with high coverage were associated with an increased risk of resistance emergence. One study simulated treatment regimens with high coverage and repeated rounds, and reported an increase in the emergence of resistant parasite strains under these conditions [57]. A study identified reduced fecundity, lower contact rate, and higher death rate as factors linked with increased resistance emergence in simulation results.

### Predictive timeline in achieving WHO’s 2020 and 2030 targets

Predictive timeline refers to the duration utilized by a modeled intervention to meet WHO targets. The definition is based on the timeframe or time horizon specified by synthesized studies for achieving either elimination or morbidity control of STH. The predictive timeline under specific scenarios is of paramount importance to enhance the policy relevance of interventions.

Synthesized model predictions with their timelines were linked to the attainment of either the WHO 2011–2020 roadmap which emphasized on morbidity control or the 2021–2030 targets with the focus on transmission interruption or elimination efforts. Morbidity control was defined as reducing moderate-to-heavy intensity (M&HI) infections to <1% in school-aged children [52], whereas the second target was the more ambitious goal of interrupting transmission or achieving elimination, defined as reducing moderate-to-heavy infection intensity to <2% in all risk groups [39]. The first roadmap served as the benchmark for the second. This shift was important for aligning with the UN Sustainable Development Goals and adopting improved WASH and community-wide treatment strategies in some settings. Achieving a target of <1% requires near-perfect WASH implementation and treatment coverage of 90%, which is extremely costly.

Through analysis of 43 modeling studies published between 2015 and 2024, 26 (60%) models simulated scenarios aligned with WHO’s 2011–2020 morbidity control targets (see Fig. 6 and table 4). Many of the modeling studies directly referenced the prevalence reduction to *<* 1% M&HI, or employed broader outcome measures such as reduction of infection intensity, morbidity control, reduction of mean worm burden, and achievement of the WHO target. Furthermore, 17 (40%) studies modeled outcomes aligned with WHO 2030 targets (see figure 6). Some explicitly mentioned a prevalence threshold of less than 2% for M&HI infection. Other terms used to describe a 2% M&HI included elimination, transmission interruption, and reduction of prevalence to nearly zero. Further, we explored whether the synthesized models specified a timeframe or time horizon (timeline) toward attaining the WHO targets. We found that only 15 studies (35%) (see Fig. 7) explicitly reported the timeline or horizon to meet either the morbidity target or elimination of STH, while 28 studies simply reported transmission interruption or morbidity control under specific conditions without mentioning the duration required for the modeled intervention to meet the defined WHO target. For example, Farrell et al. [22] highlighted that the current strategy (75% treating school-aged children) leads to the achievement of morbidity control within 5–6 years. Also, researchers in [29] reported that dual therapy reduced prevalence from 62% to 25% after 5 years, but later the reinfection rate reached 45% after stopping MDA for 39 months.

**Fig. 6:**
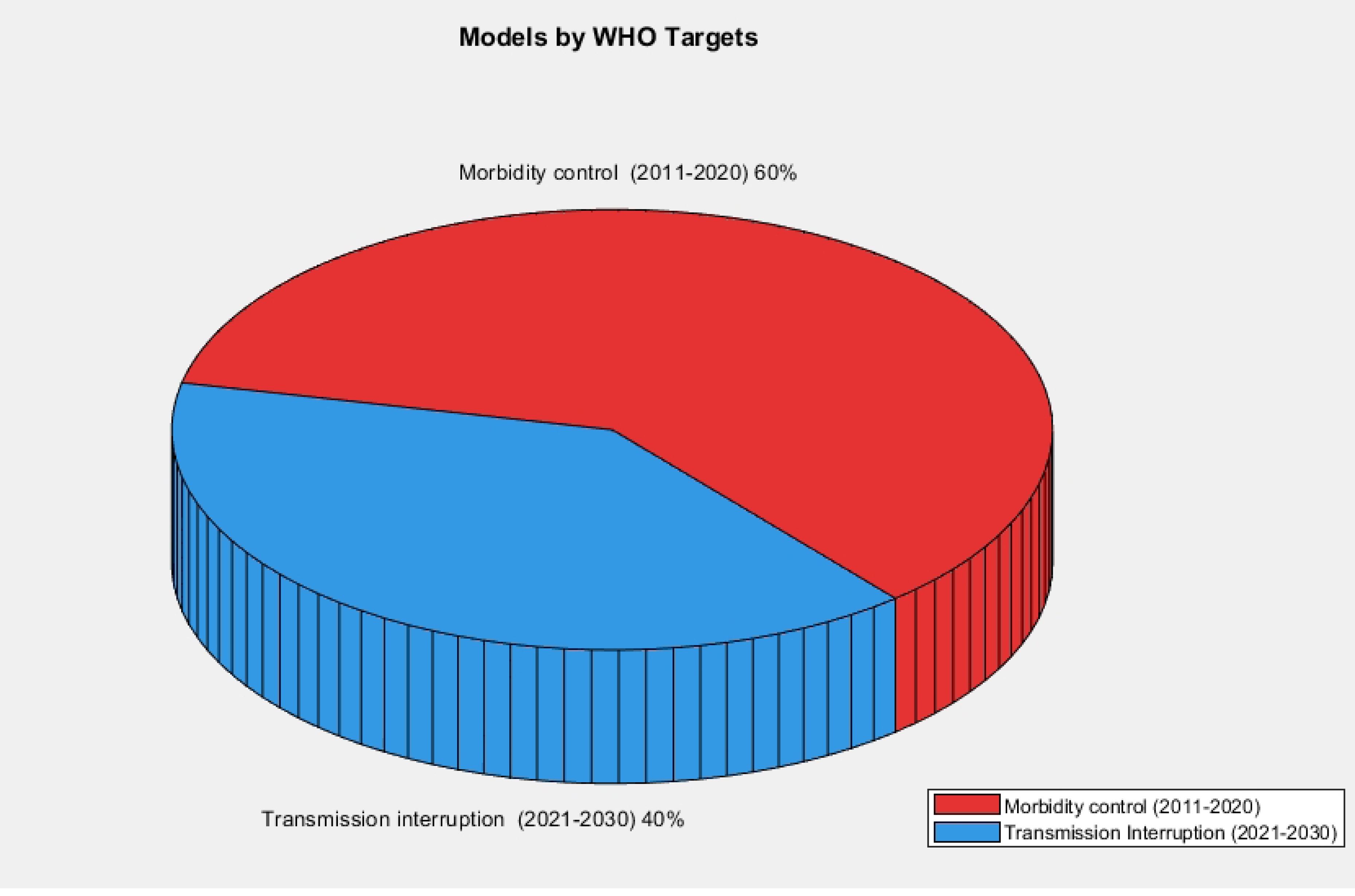
Models distribution as per WHO targets.

**Fig. 7:**
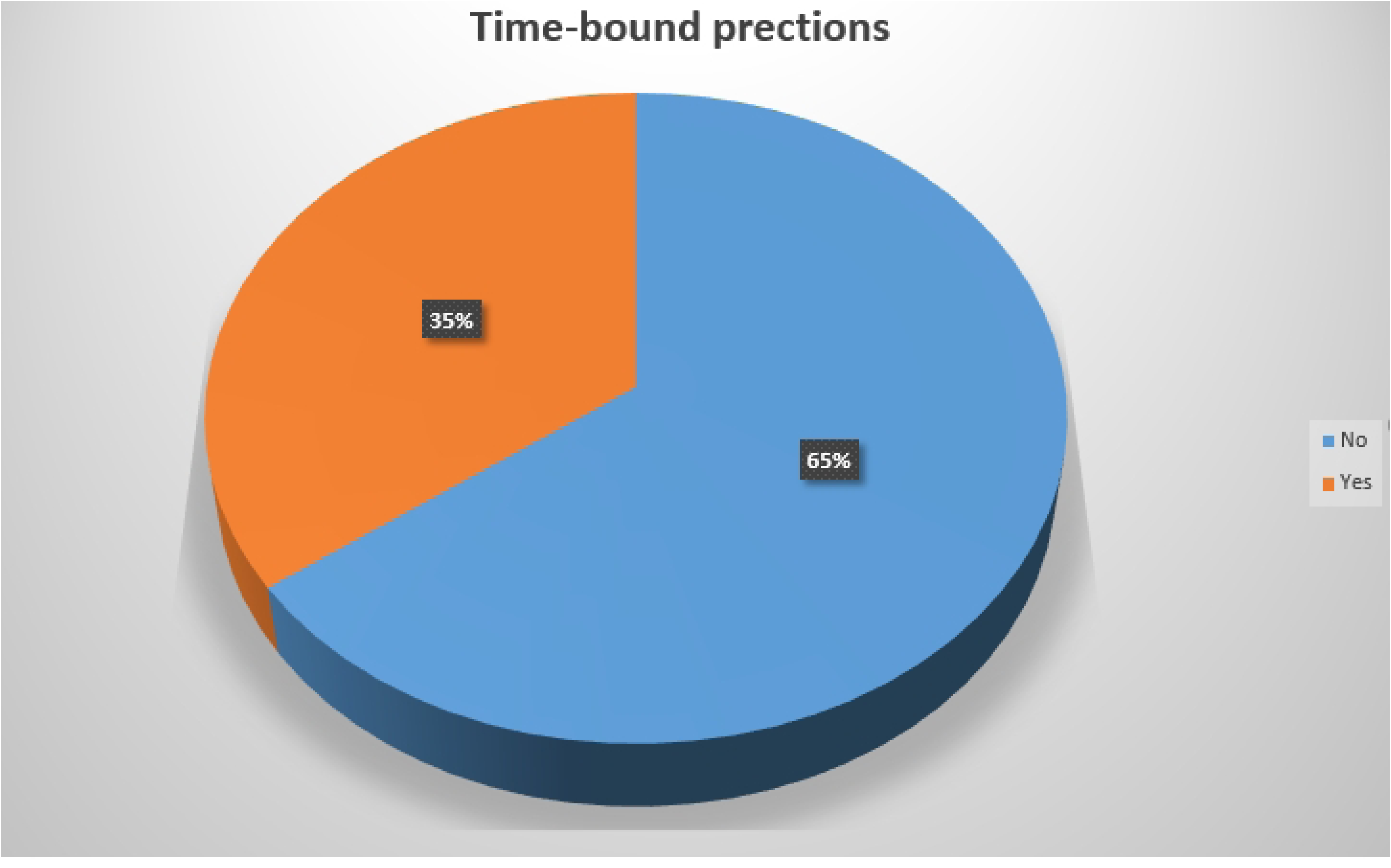
Time bound prediction reported by models.

**Table 4:** Overview of modeling studies that performed sensitivity analysis (SA) and Uncertainty Analysis (UA), highlighting their alignment with WHO targets, provision of predictive intervention outcomes, and key findings.

| Predictive Outcomes | Authors | WHO Target | SA/UA Analysis | Key Findings |
| --- | --- | --- | --- | --- |
| CBD reduces reinfection to M&HI prevalence <1% | Clarke et al. [11] | 2011-2020 | Sensitivity | Sensitive diagnostics crucial for monitoring impact |
| Higher treatment reduces mean worm burden | Coffeng et al. [15] | 2011-2020 | Both | High coverage enables elimination |
| qPCR effective for low prevalence detection | Werkman et al. [74] | 2011-2020 | Sensitivity | Prevalence $\geq 3\%$ post-MDA suggests ongoing transmission |
| WRA treatment improves child health | Vegvari et al. [71] | 2021-2030 | Not indicated | Benefits maternal/child health but may not meet targets |

| Predictive Outcomes | Authors | WHO Target | Sensitivity Analysis | Key Findings |
| --- | --- | --- | --- | --- |
| Hygiene and PC reduce $R_0 < 1$ | Mustapha et al. [41] | 2021-2030 | Not indicated | Hygiene awareness reduces infection burden |
| on-compliance increases treatment rounds | Farrell and Anderson [21] | 2011-2020 | Both | More rounds needed for long-lived parasites |
| Community MDA compensates missed PC in 2030 | Malizia et al. [35] | 2021-2030 | Sensitivity | Single community round offsets missed treatment |
| Strategy attains target in 5-6 years | Farrell et al. [22] | 2011-2020 | Sensitivity | Works for low-medium prevalence areas |
| PC + WASH accelerates elimination | Vegvari et al. [72] | 2021-2030 | Sensitivity | Biological uncertainty important for sustainability |
| Hygiene reduces infection to $< 1\%$ | Oguntolu et al. [45] | 2021-2030 | Sensitivity | Hygiene consciousness reduces burden |
| Sanitation + education cheapest | Lambura et al. [31] | 2011-2020 | Sensitivity | MDA + sanitation most effective |
| Sanitation + MDA eradicates quickly | Lambura et al. [32] | 2011-2020 | Sensitivity | Increases Mtb cases |
| Footwear + PC reduces $R_0$ | Chazuka et al. [8] | 2021-2030 | Sensitivity | Economically viable strategy |
| 80% coverage achieves elimination $< 5$ years | Okoyo et al. [48] | 2021-2030 | Sensitivity | WASH + MDA most effective |
| Community-wide reduces prevalence | Turner et al. [66] | 2011-2020 | Sensitivity | More cost-effective approach |
| WASH + PC eliminates transmission if no migration | Hardwick et al. [25] | 2011-2020 | Uncertainty | Migration affects elimination |
| High treatment risks resistance | Patel et al. [57] | 2021-2030 | Sensitivity | Faster resistance in human-infecting species |
| Prevalence reduction depends on aggregation parameter | Truscott et al. [63] | 2011-2020 | Not indicated | Non-linear prevalence-transmission relationship |
| Biannual CBD interrupts transmission at 61% coverage | Chong et al. [10] | 2021-2030 | Not indicated | Needs 61-85% coverage |
| PC reduces infection from 62% to 25% after 5 years | Kasia and Shengqiang [29] | 2011-2020 | Sensitivity | Rapid rebound after treatment stops |
| 75% MDA at population achieves target | Vegvari et al. [73] | 2011-2020 | Not indicated | 2-3% migration risks reintroduction |
| 65% PC reduces prevalence near zero within 5-6 years | Coffeng et al. [13] | 2021-2030 | Not indicated | Effective in school and community settings |
| Elimination depends on prevalence | Truscott et al. [64] | 2011-2020 | Both | Cluster size matters for elimination |
| Morbidity control can be achieved in 2 years at 75% coverage | Coffeng et al. [12] | 2011-2020 | Sensitivity | Works in high-transmission with WASH |
| Elimination possible with CBD in 10 years | Truscott et al. [62] | 2021-2030 | Uncertainty | India/Malawi success but not Benin |
| WASH uptake critical than effectiveness | Coffeng et al. [16] | 2011-2020 | Not indicated | More important than effectiveness |
| High coverage speeds resistance | Coffeng et al. [14] | 2021-2030 | Sensitivity | Intensive PC accelerates resistance |
| Morbidity attained with current program | Truscott et al. [61] | 2011-2020 | Uncertainty | Different STH responses to MDA |
| 26 infected people per year undermine elimination | Hardwick et al. [26] | 2011-2020 | Uncertainty | Stochastic models capture fade-out |
| Elimination possible for semi population wide treatments | Collyer and Anderson [17] | 2021-2030 | Uncertainty | Strongyloides knowledge gaps |
| Elimination is possible if initial prevalence is low | Chong et al. [9] | 2021-2030 | Sensitivity | Elimination vs endemic states |
| probability of elimination predicted to be in 2028 | Maddren et al. [34] | 2021-2030 | Sensitivity | Pre-SAC at risk due to low coverage |
| 75% coverage eliminates Ascaris by 2020 | Truscott et al. [60] | 2011-2020 | Not indicated | Only 22% hookworm reduction |
| Morbidity reduction depends on baseline prevalence. | Werkman et al. [75] | 2011-2020 | Sensitivity | Historical MDA affects outcomes |
| Elimination require improved WASH with PC | Okoyo et al. [47] | 2021-2030 | Sensitivity | WASH coverage critical |
| Elimination of requires hygiene consciousness | Omede et al. [51] | 2021-2030 | Sensitivity | Solutions converge to exact values |
| Elimination possible after 35 years | Turner et al. [65] | 2011-2020 | Uncertainty | Addresses albendazole limitations |
| Communtiy MDA reduced help to attain morbidity target | Turner et al. [66] | 2011-2020 | Uncertainty | Addresses albendazole limitations |
| Treatment at 90% and 80% children and adult meet elimination of worms | Werkman et al. [76] | 2011-2020 | Sensitivity | Stochastic matches deterministic |
| Seasonal timing reduces worms after 12 months of MDA cessation | Davis et al. [18] | 2011-2020 | Uncertainty | 74.5% reduction with optimal timing |
| Simple to reach less than 1% M&HI prevalence in a village level rather than a district with School MDA but easy for Community MDA | Giardina et al. [24] | 2011-2020 | Not indicated | Community MDA more effective |

| Predictive Outcomes | Authors | WHO<br>Target | Sensitivity<br>Analysis | Key Findings |
| --- | --- | --- | --- | --- |
| 88% of the village meeet<br>elimination with 90% and<br>80%coverage in children<br>and adult respectively with<br>5 years and reinfection<br>happened in 10 years | Anderson et al.<br>[4] | 2011-2020 | Uncertainty | WASH should be imple-<br>mented in the coming study |
| A significant reduction of<br>morbidity after 7 years of<br>treatment at 75% covergae<br>and 95% efficacy | Anderson et al.<br>[5] | 2011-2020 | Sensitivity | Emphasize on education<br>and WASH to the future<br>study. |

Increase of the treatment frequency improve the intervention outcome for instance biannual treatment of children and women of reproductive age reported to facilitate in morbidity attainment within 2 years [12]. Also, different helminth species have distinct timeline for example one study revealed that the morbidity target for *A. lumbricoides* is possible before 2020 under school based deworming where other helminth species go beyond 2020 [60] and suggested reinfection to happen within 5-10 years. Also, high drug efficacy (95%) and treatment coverage of 75% reported to be essential to attain the WHO 2020 goal within 7 years [5] while other study conducted by Anderson et al. [4] predicted elimination to be achieved within 5 years. Study in [67] reported elimination feasibility of STH to be possible after 35 years of school based deworming (MDA *≥*75%) in settings with *R*_0_=2. Similarly, the researchers in [75] confirmed that in high transmission settings of hookworm, administering semi-annual MDA (*≥*75%) the target attained after 10 years.

Moreover, WHO set an ambitious goal of elimination or interrupting STH transmission, for instance a study predicted elimination to be achieved in less than 5 years [47] with community wide PC at 80% coverage. In addition dual therapy predicted prevalence to be closer to zero within 5-6 years [14]. Similarly, other researchers predicted that STH will be eliminated in 2028 with CBD at a random compliance [34]. Further, occurrence of covid-19 interfered the 2030 goal and one study suggest that one year missed PC can be compensated by one round community MDA [35]. Additionally, elimination depend on helminth species since Okoyo et al. [47] demonstrated elimination of hookworm to happen within less than 5 years at 80% treatment coverage. Also, Okoyo et al. [47] predicted a duration of more than 10 years for *T. trichuris* elimination under population wide PC. Modeling work by Truscott et al. [62] predicted a prevalence in many clusters close to zero in 10 years with PC in India and Malawi at 90% and 95% chance of occurring respectively.

Most of time-bound modeling synthesized under this review focused on scenario elimination without explicitly providing time horizon (see table 4), for example higher treatment frequency enhance elimination but if the treated population confer gene of resistance then drug efficacy decline [14, 57]. Also, Chong et al. [10] revealed that semi-annual CBD at 61% can interrupt the STH transmission where for annual deworming 85% is required. Migration rate can undermine elimination prospects, for instance one study reported that 26 infected people can introduce infection even in areas with effective MDA [26].

### Results on quality assessment of included studies

By using AMS tool, the selected studies possessed the score range form 62.5-95.8% (see table 3). Only one study ranged 62.5% but many studies scored more than 70% and this imply these studies fall into a high or very high category using assessment of study quality.

Fig. 8 presents a network visualization [69] of the most common terms from the titles and abstracts of the included studies, as generated using VOSviewer. The visualization is based on a co-occurrence analysis of key terms in titles and abstracts, where the colors represent different clusters. This network visualization map contains three clusters represented by three distinct colors. Red has the highest density, reflecting frequently occurring terms, while green and blue have lower density, representing more dispersed terms. Infection, control, helminth infection, intervention, preventive chemotherapy, sanitation, and hookworm infection appeared in the high-density zone, highlighting recurring themes in the literature. These terms in the clusters support the role of modeling work in informing helminthiasis control measures.

**Fig. 8:**
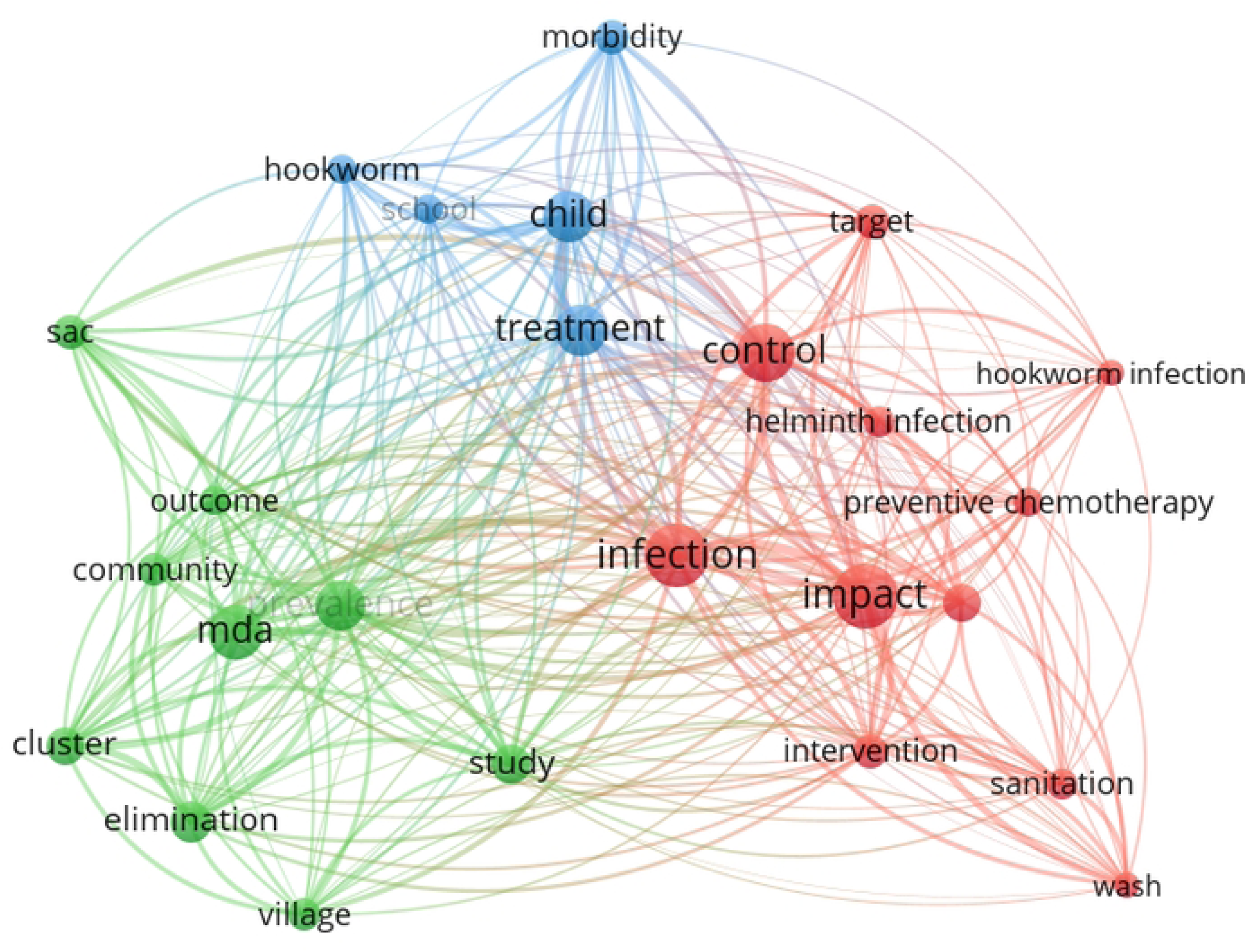
Network visualization of frequently co-occurring terms in the titles and abstracts of included studies. Warmer colors (yellow) indicate terms with higher frequency and stronger co-occurrence density.

The red cluster indicates helminth infections and targeted interventions to minimize disease impact or achieve control via preventive chemotherapy, sanitation, and WASH, as observed in the network visualization (see Fig. 8). The green cluster indicates that children are the targeted population for treatment to reduce helminth infections, though hookworm still persists. Furthermore, the blue cluster presents the focus of research studies conducted on STH, where the target population included SAC, clusters, communities, and villages. In these studies, preventive chemotherapy was administered through MDA to explore outcomes and assess elimination feasibility.

Generally visualization map was generated by determining the most frequent and thematically relevant terms with a minimum occurrence threshold of five terms.

Fig. 9 presents a density visualization obtained through text mining of common terms in the titles and abstracts of included studies. The yellow color indicates a cluster with terms evaluating control strategies such as MDA to reduce the impact of helminth infections. Terms like “elimination clusters” and “village,” as observed in the map, reflect the importance of localized interventions such as sanitation, PC, and WASH.

**Fig. 9:**
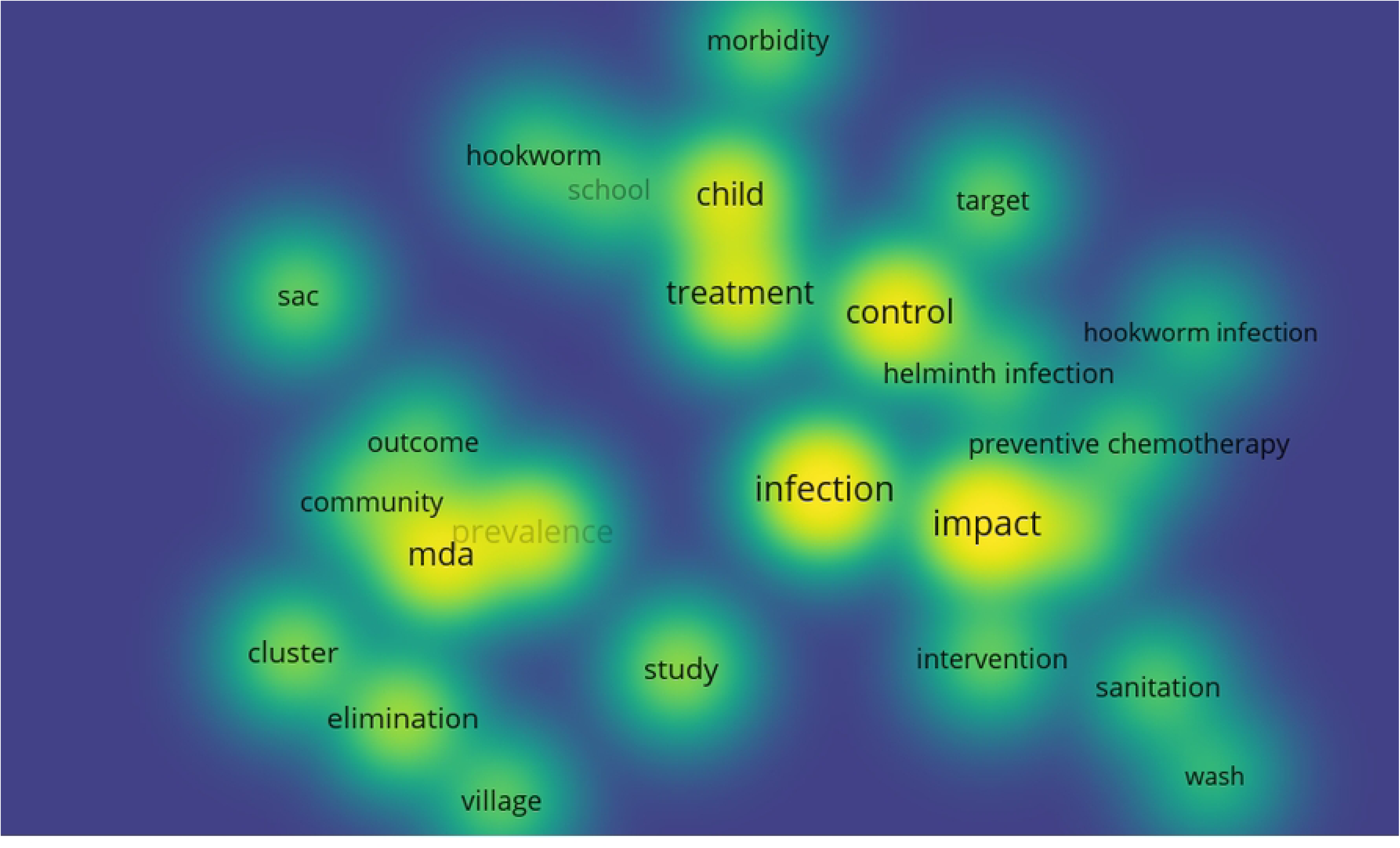
Density visualization of frequently co-occurring terms in the titles and abstracts of included studies. Warmer colors (yellow) indicate terms with higher frequency and stronger co-occurrence density.

The yellow in the density visualization map indicates that the primary risk groups for helminth infections are school-aged children (SAC). The disease impact can be controlled by distinct interventions such as MDA (preventive chemotherapy), sanitation, and WASH. The target of any intervention is to control morbidity among children, which may eventually yield significant outcomes in a cluster, village, or community under study. Centering interventions on children for helminth infection control may lead to hookworms acting as reservoirs for new infections.

Fig. 10 presents three distinct clusters of author collaborations in a connected research group. The heavy cluster (red) was centered around Anderson et al. [5], Truscott et al. [63], Hollingsworth et al. [28]. Their collaboration reflect a well established network researching on helminth modeling of control strategies. Two other prominent clusters (blue and green) featuring Farrell and Anderson [21], Hardwick et al. [25], Coffeng et al. [12], Werkman et al. [75] were collaboratively engaging in helminth infection research to provide an insight on helminth infection transmission dynamics. The network structure highlights the influence of a few central authors and the collaborative nature of the field.

**Fig. 10:**
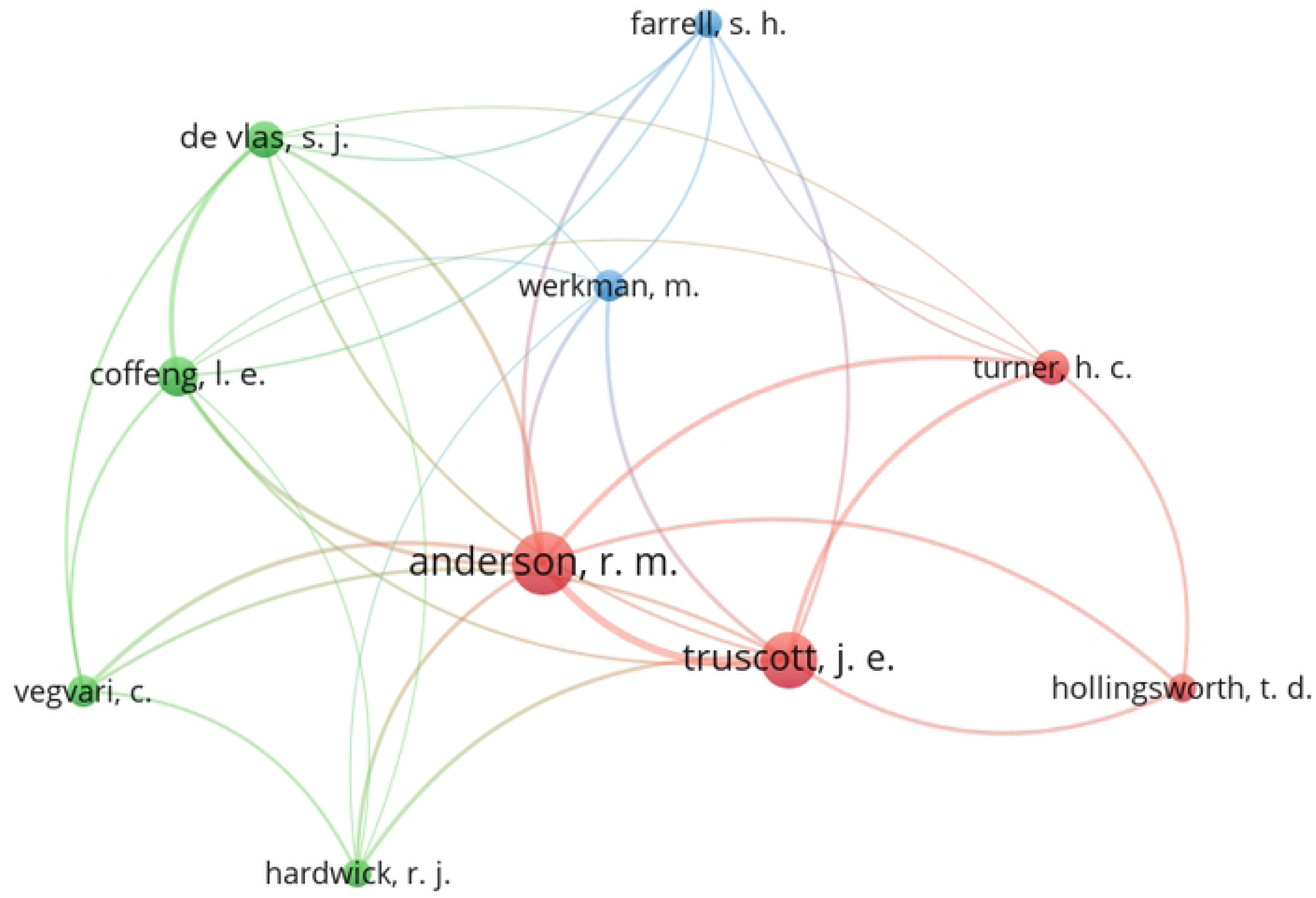
Co-authorship network visualization of authors contributing to included studies. Node size indicates publication frequency, and edge thickness represents co-authorship strength. Clusters denote closely collaborating author groups.

Much of the modeling studies employed by authors with red color are stochastic and deterministic. More prevalent models are stochastic. This made them create a strong cluster as indicated by the network visualization in the Fig. 10. Green color indicate a another cluster of authors who employed stochastic models for STH research while the blue cluster adopted an ensembe models prepared Imprerial colloge of London (ICL model) and Erasmus medical center Rotterdam (EMC models) with both having deterministic and stochastic properties.

### Sensitivity, uncertainty analysis

Developing a mathematical model of STH, in particular, depends on several pieces of evidence such as the natural history of the disease, transmission nature of the disease, and control strategy [70]. In case some of the mentioned evidence is incomplete, this leads to a sort of uncertainty in modeling. Uncertainty may be due to parameter values, methods employed, or modeling structures utilized.

Sensitivity and uncertainty analyses are essential components to enhance the credibility and applicability of mathematical models, particularly STH models. These approaches are vital in identifying the parameters that mostly influence model predictions and quantifying those predictions under different model assumptions. Out of 43 reviewed studies, 24 performed sensitivity analysis (SA), and 9 modeling studies explicitly performed uncertainty analysis (UA). Only three studies performed both SA and UA, while 5 studies did not explicitly mention either SA or UA, though this does not guarantee that they were not conducted (see Table 4 in the appendix). The selection of methods employed for SA and UA varies across studies and mainly depends on the kind of mathematical framework (deterministic, stochastic, and hybrid).

We divided the studies that performed sensitivity and uncertainty analysis into two common categories: local and global sensitivity analysis. Local SA involves changing one input while keeping other parameters fixed, whereas global SA involves interaction of multiple parameters simultaneously. Based on the synthesized studies, we further divided global SA into variance-based methods (like Fourier Amplitude Sensitivity Test) and sampling-based methods (examples include Markov Chain Monte Carlo (MCMC) and Latin Hypercube Sampling (LHS)). In total, 12 studies performed local sensitivity analysis. Twenty-four studies performed global sensitivity analysis, where only 2 considered the extended Fourier Amplitude Sensitivity Test. Additionally, eight studies implemented Latin Hypercube Sampling with partial rank correlation, while 14 performed MCMC. Across 43 reviewed studies a number of parameters were found to be most sensitive. Basic reproduction number (*R*_0_) was the most dominant parameter to inform about disease persistence or elimination. The studies in [60, 45] indicated that a small change in *R*_0_ could change outcomes, particularly in low transmission settings.

Similarly, treatment coverage, transmission rate, migration rate, and population size were sensitive parameters in many studies [62, 73, 71, 62, 47, 48].

The choice of methods for SA and UA depends on computational feasibility and analytical rigor. One-way SA is mostly used in simple deterministic models, while advanced GSA, MCMC, and others are utilized in stochastic frameworks. Nevertheless, few studies integrated both SA and UA in a cohesive framework, often due to methodological or computational constraints. For example Patel et al. [57] conducted ecological and evolutionary sensitivity analyses where pairwise analysis was done on the ecological model side and three parameters on the other models, while species-specific parameters were kept constant. Additionally, seven studies [76, 62, 12, 22, 11, 63, 24] conducted varied sensitivity analyses on detection tools like KK, PCR, and qPCR.

### Model validation

Credibility and predictive power of the model depend on validation. The reviewed studies employed both insample (internal) and out-of-sample (external) validation processes to enhance model utility. Many modeling studies checked for internal consistency by comparing model output with the same datasets used in calibration [70].

In-sample validation involves examining how well the model fits the same datasets utilized to build or calibrate the model. It is a validation against past data or historical trends to evaluate alignment between predicted and observed values. In-sample validation is commonly referred to as model fitting or calibration. It also involves optimizing model parameters to minimize the discrepancies between predictions and observations in order to improve forecasting of intervention impact and effectiveness. In total, 30 studies conducted in-sample validation. Among these, 11 studies employed Bayesian techniques such as MCMC and sequential Monte Carlo for validation. Statistical methods such as non-linear least squares techniques and maximum likelihood estimation (MLE) validated 16 modeling studies. Only 3 studies adopted both Bayesian and statistical approaches when conducting internal validation. The goodness-of-fit metrics included visual comparison of simulated against observed data, residual analysis, and model comparison approaches such as Bayesian Information Criterion (BIC) and Akaike Information Criterion (AIC).

The reviewed studies employed a wide range of model fitting techniques, ranging from statistical methods to Bayesian estimation. Maximum Likelihood Estimation (MLE) was widely used to fit parameters such as drug efficacy, diagnostic sensitivity, and transmission rate, particularly in deterministic frameworks (see Table 2) [60, 64, 63, 62, 66]. Additionally, MLE was used to estimate transmission rate and *R*_0_ by fitting the model to cross-sectional individual-level data from St. Lucia [6], which included age-specific egg counts and worm burdens [25]. Least squares techniques were employed on time series data to minimize discrepancies between model predictions and observed intensity via nonlinear regression [41]. Bayesian estimation methods such as Approximate Bayesian Computation and Bayesian inference were employed to account for uncertainty and parameter distribution [18, 17, 15]. These approaches use prior knowledge and observed data for estimation of model parameter posterior distributions.

Approximate Bayesian Computation employed to fit parameters to age-stratified prevalence data from Cambodia [17]. Coffeng et al. [15] employed the ICL/EMC model, which was previously validated using Maximum Likelihood Estimation, while the validation approach for the EMC model was Bayesian Sequential Monte Carlo (SMC). Validation was done using two approaches; a qualitative comparison of model predictions with observed patterns of reinfection past treatment was one form of validation, where observed reinfection after several rounds of intervention was compared with expected patterns [60]. Also, MCMC was employed to estimate *R*_0_, egg production, and contact parameters for three age groupings [60].

Out-of sample validation was rare in the synthesized studies. External validation enhances model generalizability. These validated models may inform whether the 2030 WHO target can be attained using available control measures regardless of mode diversity in different settings. Only 4 [62, 29, 60, 71] modeling work compared model prediction with observation data other than one used for model fitting. The mentioned studies conducted both internal and external validation. For example study by Kasia and Shengqiang [29] explicitly conducted external validation by fitting their hookworm transmission model to independent field datasets from multiple geographic locations (China and Zimbabwe) and comparing model predictions with observed prevalence trends under different chemotherapy programs in three distinct location. Also, the researchers in [60] model was validated against longitudinal reinfection data from [19, 20], which recorded worm burdens pre- and post-treatment in a South Indian community after that they compared with independent datasets.

## 4 Discussion

Mathematical models are an indispensable tool for understanding disease transmission dynamics and guiding the evaluation of strategies to control morbidity or eliminate infections in public health [36]. The study at hand synthesized evidence from 43 mathematical modeling studies that evaluated STH transmission dynamics and control strategies. The key findings address model types based on structural assumptions and population features, predictive intervention effectiveness for achieving WHO targets, and the timeframe required to reach WHO goals for NTDs, particularly STH.

Deterministic frameworks, representing 44% of the reviewed studies, are traditional frameworks employed to understand mechanistic processes, such as how certain interventions affect *R*_0_. These models are essential for parameters sweeps and evaluating the effectiveness of a single intervention in the modeling process [31, 45, 8]. Further, the these frameworks are computationally inexpensive and analytically tractable. Deterministic frameworks remain significant in the initial exploration and evaluation of interventions at the population level when data are scarce, but they fail to capture heterogeneity in PC uptake and exposure to infection at the individual level [14]. The relatively lower number of deterministic frameworks indicates a shift to more powerful models for better disease prediction.

Stochastic models (47%) sightly outnumbered the deterministic approaches. The growing application of stochastic models reflects an earlier shift necessitated by the need for complex models that capture random fluctuations, uncertainty, and low prevalence or elimination feasibility of STH. The modeling transition supports the 2030 WHO goal of interrupting STH transmission or eliminating it as a public health problem. For example, Clarke et al. [11] and Werkman et al. [75] demonstrated community wide treatment to be better in elimination of STH compared to child treatment strategy. Moreover, stochastic frameworks require moderate to high computational power, and their analytical analysis is often infeasible necessitating extensive simulation and probabilistic interpretation.

Hybrid models, accounting 9% of the synthesized studies, act as the middle ground between deterministic and stochastic modeling studies [1]. The relatively low number of hybrid frameworks reflects their technical complexity, though they present an opportunity for future research to focus on balancing realism and analytical tractability. Hybrid models are complex since they employ both aggregated trends and scaled variability, yet they are powerful for evaluating elimination of STH near breakpoints [25]. In addition these multi-scale frameworks are suitable for migration dynamics investigation for instance researchers in [26] employed a hybrid approach to demonstrate that even 26 infected migrants per year can reintroduce helminth infection in an area with effective MDA. Similarly hybrid models are powerful frameworks that capture both randomness and diverse population characteristics, making them particularly strong for heterogeneity investigations where stochastic models fail to represent population diversity. Moreover, hybrid models can handle both randomness and diverse population structures—making them more comprehensive for capturing heterogeneity.

Models were further classified into three sub-classes based on population representation and heterogeneity in space with age-structured models comprising majority (56%) of the population-stratified frameworks. These models capture differences in susceptibility, transmission dynamics and demographic processes such birth and mortality rates as across age groups [47]. The dominance of age-structured frameworks reflects the biological and epidemiological importance of age in helminthiasis transmission dynamics and burden, as all three parasite species show strong age-dependence. School-aged children exhibit high prevalence and intensity due to behavioral and immunological factors. Such modeling enables age-specific or cohort-specific intervention assessment, including: school-based programs [67], targeted deworming [71] population-level treatment [66, 22]

The availability of age-specific prevalence data may further explain the predominance of age-structured models. Age-structured stochastic models prove particularly essential for STH elimination targets, as multiple synthesized studies reported community deworming’s superior effective compared to school-based approaches. Regarding species-specific dynamics, studies revealed that Ascariasis and Trichuriasis are most prevalent in children, while hookworm primarily affects adults. Age-structured models evaluated species-specific interventions, with several studies reporting that *A. lumbricoides* responds faster to benzimidazoles than hookworm and *T. trichiura*. These structured models also demonstrated that treating high-risk groups like pregnant or lactating women improves child health by reducing anemia and increasing birth weight [71].

The modeling study by Coffeng et al. [12] highlighted a linear relationship between hookworm intensity and age (increasing from children to adults), whereas *A. lumbricoides* and *T. trichiura* infections peak in childhood and decline in adulthood age. Collectively, the reviewed age-structured modeling transformed STH control by: revealing limitations of school-based deworming for hookworm, quantifying the role of adult reservoirs and informing WHO’s age-extended treatment recommendations.

Flat models, representing 28% of the reviewed studies, were utilized in scenarios where age stratification was unnecessary due to limited data availability. These models often serve as foundational frameworks for initial exploration, assuming homogeneous population mixing in closed environments. While uniform mixing facilitates intervention testing, it oversimplifies transmission heterogeneity and may yield inaccurate STH elimination predictions. Closed population assumptions also neglect reinfection risks from human mobility between connected localities, making these models unsuitable for localized outbreak analyses. However, non-structured frameworks remain valuable for large-scale intervention policymaking due to their analytical and numerical cost-efficiency. Their flexibility allows extensions to incorporate: co-infection dynamics, age structure or spatial patches, environmental drivers, drug resistance compartments [32]

Spatial models, accounting 16% of the reviewed articles were relatively lower. However, these frameworks add an element of realism due to the incorporation of geography, network interaction, and movement [73]. They were employed where populations or infections are not uniform in space. Similarly, spatial models allow for dispersal (clustering villages) and human movement. Moreover, it is possible to explore geographical interventions under these frameworks. In addition they are flexible for mobility and connectivity incorporation; for example, metapopulation models allow investigation of disease spread between villages, households, rural or urban areas, and cities via travel routes [62, 72]. Furthermore, these models are suitable when interactions depend on proximity, systems are location-dependent, and when explicit spatial data are available. However, these models are harder to analyze analytically.

Spatial models demand intensive geo-referenced data for parameterization; therefore, lack of such data reduces the number of these models in the field. Growing literature on spatial models indicates that STH studies focus on hotspot-targeted interventions and the impact of human movement. Additionally, they capture heterogeneity in terms of host movement, control measures, multi-helminth species mix, and inter-patient variation in the duration of infection and virulence [2, 3]. Similarly, these models are important in reflecting STH control in a local context by examining localized outbreaks but are computationally expensive.

A major trend observed across the synthesized studies is the dominance of comparison between school based and community wide deworming. 18 studies contrasted the two interventions within one modeling framework. The dominance of this comparison reflects ongoing debates on which intervention accelerates progress toward achieving WHO targets. The two control strategies were contrasted to determine which yields superior outcomes in achieving WHO targets of eliminating STH or controlling their morbidity. SBD intervention was reported to achieve the 2011–2020 WHO targets in school children but is limited in achieving the elimination goal. Further, biannual treatment was reported to be better than annual, and high treatment coverage improves outcomes. To achieve the elimination goal, SBD should be used as a baseline intervention. Generally, SBD is logistically simpler and more cost-effective for SAC, but CBD is more impactful for community-wide elimination, especially in high-endemicity settings [66].

A number of studies reported CBD to be superior in interrupting STH transmission compared to SBD [66, 21, 65, 35, 64, 63, 10]. CBD is highlighted as strong intervention in achieving WHO 2021–2030 targets and has a shorter timeline to impact. The intervention is characterized by equitable protection since it treats almost all people. It is cost-effective over time in achieving elimination goals and reducing reinfection but is logistically intensive. CBD requires higher resources and broader infrastructure, making implementation challenging in low-resource areas.

8 studies modeled PC without specifying delivery platforms such as school-based, community, or adolescent girls. The presence of studies modeling PC indicates that the intervention remains the backbone but has considerable limitations like resurgence effects. Also, the decrease in studies modeling PC alone may indicate that the world is now focusing on testing integrated control approaches. High PC coverage is highlighted to reduce STH effectively. Integrated approaches with WASH and PC shorten the elimination timeline [47]. WASH helps reduce environmental contamination and reinfection. There is an increase in the number of studies incorporating WASH with PC (10 studies), implying that PC alone cannot reach elimination in supporting 2030 targets. Different helminths respond differently to this integrated approach; for instance, *A. limbricoides* responds highly to MDA and WASH intervention due to its low lifespan [66], but hookworm requires adult-targeted MDA, and *T. trichiura* demands alternative control strategies [61]. Also, data availability may increase more modeling on integrative control approach of WASH and PC.

Benzimidazole derivatives have low efficacy against hookworm and *T. trichiura*, implying the need for alternative strategies or dual drug therapy [66]. Combination therapy has been reported to improve cure rates and egg reduction rates, especially for *T. trichiura*. Only three studies employed dual therapy [29, 65, 13]. Co-administration of albendazole with ivermectin raised the egg reduction rate to within the range of 92–95% for *T. trichiura* compared to 60% with monotherapy [65]. Dual therapy was highlighted to be effective and can delay drug resistance development [57, 14]. Comparative models from the Erasmus Medical Center of Rotterdam (EMC) and Imperial College London (ICL) predicted improvement by co-administration of albendazole and ivermectin rather than albendazole monotherapy.

Also, fewer models targeted women of reproductive age and adolescent girls, indicating a gap in gender-responsive modeling despite WHO calls for gender-inclusive modeling. Targeted deworming of SAC and WRA (2 studies) modeled together demonstrated improvement in disease outcomes, particularly reducing anemia to enhance better health in children. The lower modeling on hygiene (1 study) and school-based deworming (1 study) may be due to a shift from modeling standalone interventions with slower impact trajectories to integrated control approaches with higher effectiveness in controlling STH.

The synthesized modeling studies of STH control published between 2015 and 2024 captured a shift from morbidity control to transmission interruption or elimination as stipulated by the WHO’s 2011–2020 and 2021–2030 NTD roadmaps. The analysis reveals a stronger alignment (60%) with the morbidity control targets (2011–2020 NTD roadmap) compared with the elimination-focused goal (2021–2030 NTD roadmap). This indicates that the earlier synthesized mathematical models focused on quantifying the impact of preventive chemotherapy (PC) for attaining morbidity control targets in school-aged children. The relatively lower number (40%) of modeling studies aligned with the 2030 WHO goal may be due to the recency of the target, as based on the review duration (2015–2024), only articles from the last four years were included. Also, the lower publication rate may be due to the complexity involved in transmission interruption or elimination dynamics.

Moreover, out of 43 studies, 15 (35%) frameworks explicitly reported the timeline required for the intended WHO goal. Further, only 6 studies [47, 14, 62, 34, 35, 72] provided a timeframe for achieving STH transmission interruption or elimination feasibility (2021–2030 WHO targets). Studies in the literature described models’ output in relative terms such as prevalence reduction or elimination of the disease without mentioning the exact time for reaching the mentioned targets (see table 4). The limitation of reporting timelines in the reviewed studies reflects the complexity of the models since they may be required to capture many factors for elimination such as environmental transmission, WASH, population-wide intervention, and reinfection data, which may be insufficient. Many studies focused on comparing the effectiveness of a strategy rather than reporting a timeframe. In addition, the number of studies reported the time required to achieve morbidity targets was relatively high due to the simpler and shorter term of the goal. The lack of time-bound prediction limits model utility for policy planning, specifically on confidently reporting whether the modeled intervention will explicitly meet the target within a specific duration. A realistic time horizon is useful in planning national deworming programs and tracking progress, and it is essential due to scarcity of resources. Reporting timelines should be a standard practice for models with the intention to support global health targets, in particular STH elimination models.

### Strength and limitation

The study at hand benefits from a comprehensive literature search strategy, which in turn yielded a wide array of mathematical models on STH control. The study employed Web of Science, Embase, and Scopus databases rich in peer-reviewed literature from epidemiological and mathematical disciplines, ensuring relevance to STH control studies. However, using only three databases may have reduced the search query outputs.

This overview relied on results obtained from extensive literature search from databases by using a search term describing transmission dynamics mathematical models that evaluated control strategies. The evaluated models incorporated different intervention strategies to either morbidity control or interrupting STH transmission. Notably, this study is the first to systematically synthesize whether mathematical models explicitly predicted the time horizon required for modeled interventions to meet WHO morbidity control or interrupt STH transmission - an essential but often overlooked aspect in planning national deworming programs, tracking progress, and evaluation.

The search strategy was built based on common terms used in peer-reviewed literature, although the search term yield a significant number of articles, it may not be exhaustive in capturing all relevant papers. Also, the study included only English articles as per the authors’ first language. Furthermore, systematic reviews, book sections, press articles, pre-print materials, letters to editors, and conference proceedings were excluded as they did not meet the inclusion criteria requirements, yet excluding of this grey literature may lead to missing of good information to shape the study. Fig 2 shows the published literature, but the articles are not evenly distributed yearly, with no single identified article in 2023. The study considered only transmission dynamics models that evaluated interventions. Yet, statistical models, Bayesian approaches, and machine learning models might have impacted the review results, but these studies were excluded. Further, the review team was trained to identify mathematical models of transmission dynamics that evaluated control strategies. However, there is a possibility that some important articles were excluded due to reviewers bias. We limited the review to the past 10 years for manageability and to ensure the inclusion of recent and relevant research. Although the review assessed how studies reported progress toward two WHO targets on NTDs, the search duration from 2015 to 2024 excluded articles published between 2010 and 2014 which may have provided valuable insights into the 2010–2020 WHO target. Additionally, articles published between 2025 and 2030 may offer more substantial contributions toward evaluating the 2030 WHO target. Moreover, parts of the AMS tool used to appraise the reviewed articles relied on the reviewers’ subject knowledge, which may introduce potential bias.

Similarly, there is no a study in the literature has modeled long-term transitions between SBD and CBD, and this gap limits policymakers’ ability to predict outcomes of programmatic shifts. Additionally, the underrepresentation of models targeting women of reproductive age (WRA) and adolescent girls highlights a critical gap in gender-inclusive modeling, despite WHO’s emphasis on equitable and inclusive control strategies.

## 5 Conclusion

The present study synthesized the published landscape of mathematical models describing the transmission dynamics of soil-transmitted helminthiasis (STH). The study compared structural assumptions across models, assessed the effectiveness of various control strategies in achieving WHO targets, and examined the extent to which models provided time-bound predictions for reaching morbidity control and transmission interruption. This review represents one of the first comprehensive efforts to critically appraise the capacity of STH models to forecast timelines for achieving programmatic goals.

The synthesized studies revealed that stochastic models (47%) were slightly more prevalent than deterministic models (44%), with relatively few studies employing hybrid frameworks (9%). The increasing preference for stochastic models reflects a shift from earlier modeling research that predominantly favored traditional deterministic approaches to the complex and realism frameworks which support rare events like STH elimination. Such modeling approaches align with the 2030 WHO goal of interrupting STH transmission.

Furthermore, in terms of population stratification, age-structured models (56%) were more prevalent compared to non-structured (28%) and spatially explicit frameworks (16%). The dominance of age-structured models reflects the biological and epidemiological relevance of age in helminth transmission dynamics and disease burden, as the three main species of soil-transmitted helminths (STHs) exhibit strong age-dependent patterns. Such models enable the assessment of age-specific interventions such as school-based deworming, targeted treatment of high-risk groups, and broader population-level interventions.

Among the reviewed studies, school-based deworming (SBD) and community-based deworming (CBD) were the most commonly compared interventions, featured in 18 modeling studies, followed by integrated strategies combining PC with water, sanitation, and hygiene (WASH). Community wide treatment confirmed to be consistently effective to interrupt STH transmission in most settings. Similarly, integrated approaches offer more sustainable and impactful interventions compared to standalone strategies.

Only 15 of the 43 reviewed studies explicitly highlighted the expected time horizon for achieving either morbidity control or elimination of STH under specific conditions. The timelines varied significantly across studies where their range was between 2 and 35 years

### Recommendations

Based on this study’s findings, we recommend the following research on STH control. Future research should focus on modeling that: explores the impact of multi-drug combinations such as triple therapies; integrate PC and WASH interventions; extend models to capture co-interactions between STH and other pathogens like malaria, Mycobacterium tuberculosis, and other neglected diseases; capture heterogeneity of disease by using either spatial or hybrid models; consider seasonality to capture environmental variables; simulate the switches between SBD and CBD while varying time and threshold parameters; explicitly reporting on intervention timelines to reach WHO targets; model realistic constraints such as treatment fatigue, resurgence risk, and sub-optimal coverage to improve model applicability; and account for drug resistance development.

The findings from this review have direct implications for policymakers, program implementers, and funding agencies involved in STH control: policymakers should transition to community-wide deworming, as modeling evidence demonstrates it to be more effective in almost all transmission settings compared to school-based deworming, particularly for achieving 2030 targets. Also, to enhance STH strategic planning, policymakers should support modeling studies that explicitly report the timeframes required for interventions to achieve either morbidity control or transmission interruption. This is essential for phased intervention implementation and funding cycles. National STH policies should promote integrated control approaches such as WASH complementing PC and dual therapy when drug efficacy is low. This will enhance the impact and effectiveness of interventions toward achieving sustainable control of STH. Validation improves confidence in model predictions and their utility to guide STH control, so policymakers should support the development and validation of modeling studies with real data. Research agencies and policymakers should promote the development of hybrid and spatial models to capture real complexities in geography and heterogeneity in transmission coverage.

Conclusively, mathematical models are essential tools in shaping STH control strategies and inform on WHO targets. The move to 2030 goals of STH elimination or interrupting its transmission, models must evolve to report on effectiveness of intervention, and timeframes required for that intervention to meet 2030 goal. The study improves understand for effectiveness of current control strategies towards attaining WHO 2030 goal and future research effort should be on models with integrative, time-bound predictions, policymakers frameworks shaping achievement of 2030 targets.

## Supporting information

### S1 PRISMA Checklist

PRISMA guidelines for full text of review and abstract. (DOCX)

## Authors’ contributions

Lemjini Masandawa conceived the study. Lemjini Masandawa, Safari Kinung’hi, and Isambi Mbalawata design the methodology. Miracle Amadi, Silas Mirau and Lemjini Masandawa did data extraction and analysis. Miracle Amadi and Silas Mirau performed interpretation. Lemjini Masandawa and Miracle Amadi drafted the manuscript. Silas Mirau and Safari Kinung’hi were project administrator. All authors read and approved the final version of the manuscript

## Data Availability

no data

http://www.example.coma2

## Acknowledgments

The authors acknowledge with thanks the support of all authors

